# Transmission Networks and Intervention Effects From SARS-CoV-2 Genomic and Social Network Data in Denmark

**DOI:** 10.64898/2026.01.08.26343683

**Authors:** Jacob Curran-Sebastian, Christian Morgenstern, Jonas Juul, Mark P. Khurana, Neil Scheidwasser, Nicolas Banholzer, Alexandros Katsiferis, Frederik T. Møller, The Danish COVID-19 Genome Consortium (DCGC), Pikka Jokelainen, Jannik Fonager, Anders Hviid, Morten Rasmussen, Marc Stegger, Sune Lehmann, Tyra G. Krause, Laust H. Mortensen, David Duchene, Neil M Ferguson, Samir Bhatt

## Abstract

Decision-making by governments during disease outbreaks is increasingly reliant on large-scale pathogen genomics and detailed individual-level data. We use 293,841 SARS-CoV-2 genomes collected in Denmark, between September 1st 2020 and December 31st 2021, and combine these with comprehensive individual-level data on settings, including households, schools, workplaces and family relationships, to infer plausible transmission pathways. Next, we use the plausible transmission pathways to assess the effectiveness of specific non-pharmaceutical interventions (NPIs) and quantify transmission heterogeneities, providing a more detailed understanding than is possible from aggregate national estimates. School restrictions were associated with reduced community transmission; face coverings and gathering restrictions were associated with reduced transmission across all settings. We find that those vaccinated with two doses, compared to the unvaccinated, had onward transmission reduced by 23.5% (95% highest posterior density interval: 21.5-25.2%). Our proposed approach is pathogen agnostic, and can be used in future outbreaks where genomic data and data on social relationships are available.

## Introduction

Pathogen genomic surveillance enables the reconstruction of transmission pathways with fine granularity, revealing patterns of spread, variability in transmission dynamics, and the effects of non-pharmaceutical interventions. However, population-level transmission analysis remains an enormous challenge, and many existing methods, particularly in Bayesian inference [1, 2], do not scale well beyond a few hundred genomes and are unsuitable for large-scale genomic surveillance of infectious diseases.

Our response to pandemics is likely to be dramatically improved by the use of multimodal data on sample networks and settings [3], allowing accurate reconstructions of transmission trees [2, 4]. In this study, we use 293,841 SARS-CoV-2 genome sequences collected between 1st September 2020 and 31st December 2021, a period of intensive testing and sequencing during which 37.6% of all positive polymerase chain reaction (PCR) tests were sequenced [5], rising to 81.9% between 1st February and 1st October 2021. This sequence data is linked to extensive data from the Danish registries and national-scale social network data, which details the social relations (household, school, workplace and family relations) of every individual living in Denmark [6]. We develop a novel framework that is both fast and scalable to derive plausible transmission networks from this linked dataset.

Informing non-pharmaceutical interventions (NPIs) is a crucial outcome of transmission analysis, with NPI effectiveness usually estimated using aggregate data on their variation and stringency across countries [7–10]. However, national-level models using aggregate-level data mask local heterogeneity in NPIs, transmission dynamics, and sociodemographic context, risking ecological fallacies and potentially biased effect estimates [11]. Studies that use data at the individual level provide a more detailed and nuanced understanding of how interventions affect transmission across different settings, thereby accounting for sociodemographic heterogeneity [12–14]. Here, we assess the effectiveness of NPIs using individual-level data and quantify variation in transmission risk across and between individuals to estimate, for example, the impact of vaccination on SARS-CoV-2 transmission. Whilst comprehensive contact surveys may be utilised to investigate infection contact networks [15], as done in China early in the SARS-CoV-2 pandemic [16], our framework enables us to characterise transmission dynamics at the national scale for Denmark over a prolonged period, significantly improving our insights into interventions and risk heterogeneities.

We identify plausible transmission pairs using a mechanistic model of between-host evolution that scales efficiently to the 293,841 genomes in our dataset, and integrate these results with individual-level data to reconstruct potential transmission networks. Despite the scalability of our approach and its applicability to similarly large genomic datasets, we highlight that the resulting networks depend critically on the inclusion of other data sources, particularly information on social relationships. We show how combining these data sources can give insights into transmission pathways and the impact of interventions, including school closures, vaccination, and stay-at-home orders (an outline of our analysis is given in Figure 1).

**Figure 1.**
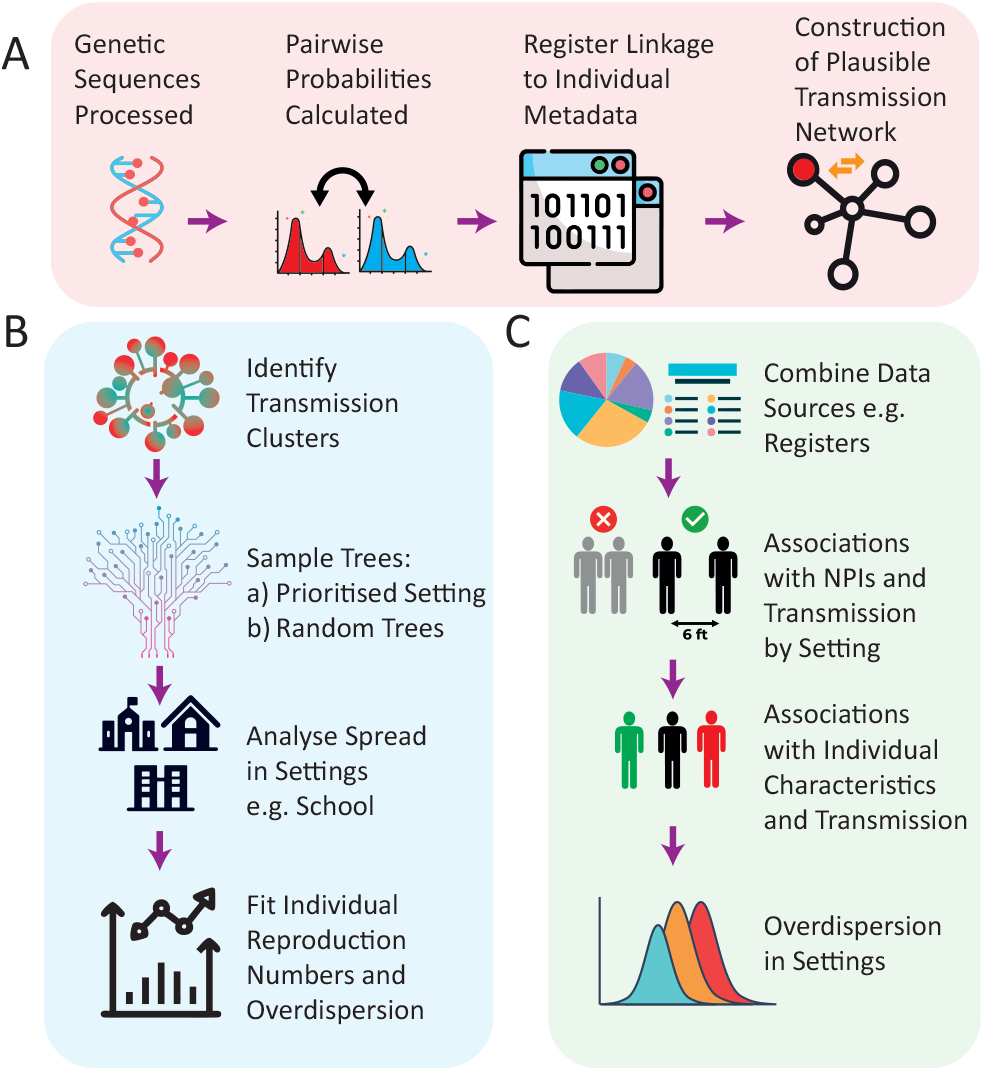
Schematic outline of the paper: (A) Construction of plausible transmission networks based on genomic sequences, testing dates, and registry data, (B) identification of transmission clusters, trees, transmissions using social network data to assign settings, and inferred reproduction numbers, and (C) using transmission networks to assess setting-specific NPI effectiveness based on individual-level data and risk heterogeneities for transmission.

## Results

### Construction of Transmission Networks

We combine genetic sequence and testing data to construct a directed network, *G*, of *plausible transmission pairs*. We add edges between individuals based on a mechanistic Poisson process model of viral evolution assuming a substitution rate of 0.64 substitutions per week (similar to e.g. [17]) and a generation time following a Gamma(4.87, 1.98) distribution, whose parameters are the mean and variance of the distribution [18] (see Methods for details of this model). We then define two individuals *A* and *B* to be a plausible transmission pair, with *A* a plausible infector of *B* (drawing an edge in *G* from *A* to *B*), if their genomic sequences differ by at most 2 nucleotide substitutions, and if *A* was tested positive before *B* within at most 11 days (see Supplementary Figure 5 for the full range of values that define a plausible transmission pair).

From this network, we sample 100 plausible transmission trees by choosing at random a single infector for each individual from their set of plausible infectors. For our main analysis, we prioritise choosing infectors who share one of the settings defined by the nation-scale social network with their plausible infectee (which we refer to as “prioritised settings”). As a sensitivity analysis, we compare these results with trees in which infectors for each individual are sampled at random from their set of plausible infectors, regardless of which settings they occupy (referred to as “random trees”, see Supplementary Figure 7).

Figure 2A shows the weekly proportion of sampled individuals that have a plausible infector in one of the settings identified by the nation-scale social network, which offers information on household, school, workplace and family relations for every individual living in Denmark (see Methods and [6]). Using our model, we find that a weekly proportion of up to 56% of sampled individuals have a plausible infector belonging to either their household, school, workplace or family. Over the study period, we find that a total of 35.7% of sampled individuals have a plausible infector that shares one of these settings. Some of the remaining proportion of individuals without a plausible infector in one of these settings could be explained by the under-ascertainment of cases that are not identified through testing, although we estimate that case ascertainment is consistently high throughout much of the study period (see Supplementary Figure 1). In Supplementary Figure 7, we show that the “random trees” sampling procedure results in a maximum of 13.7% of weekly transmission attributed to settings, which is inconsistent with literature estimates [16, 19, 20], highlighting the importance of including social network data in our “prioritised settings” approach.

**Figure 2.**
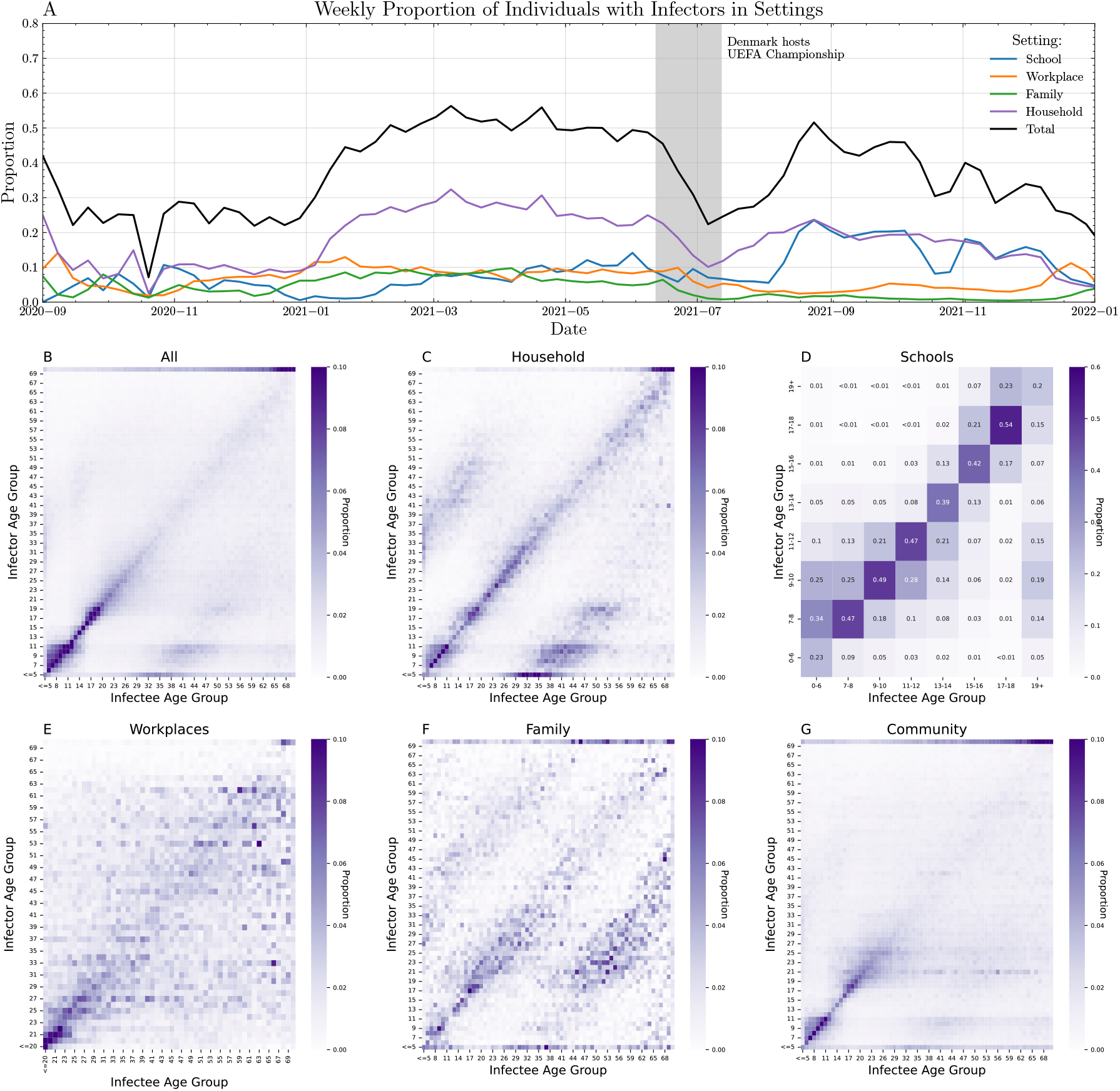
(A) Weekly proportion of individuals whose sample infector belongs to different settings for each variant for 100 random sampled trees with settings-based transmission prioritised (“prioritised settings”). The total proportion (black line) is the sum of the proportions of individuals across all settings (households, schools, workplaces and family) (B-G) Age-structured matrices showing the proportions of people in each setting with sampled infectors belonging to different age groups. Panel B contains all sampled infector-infectee pairs regardless of setting, and panel G contains sampled community infector-infectee pairs, who do not share any of the settings defined in the nation-scale social network. Proportions shown represent the median values across 100 sampled trees.

Of our defined settings, we find that plausible household pairs consistently represent the highest proportion of plausible transmissions (up to 30%), followed by schools and workplaces. Supplementary Figure 2 shows the testing and sequencing proportions of individuals in different settings and age groups. We identify a drop in July 2021 to 22.4% of individuals with a plausible infector in settings; however, this drop coincides with a large outbreak linked to the hosting of the UEFA European Football Championships by Denmark in that same period [21], with many infections occurring amongst attendees of the tournament. We also observe a noticeable drop in transmission within schools during the final week of October in 2021, coinciding with school closures for the autumn holiday celebrated in Denmark, during a period of otherwise high transmission within schools.

Figure 2B-E show heatmaps of the age distribution of transmission pairs in each of the four settings identified in the nation-scale network from our sampled transmission trees. Across all settings, the age structure matrices display the expected strong main diagonal, reflecting frequent interactions among peers of similar age, and resembling patterns found in other studies [15, 22] but estimated from observational data and not contact surveys. However, each setting also reveals distinct secondary patterns. Households closely resemble the classic contact matrix structure previously reported [15], where in addition to peer interactions, pronounced child–parent contacts are evident. Family networks broaden these intergenerational connections beyond the nuclear household, producing multiple off-diagonals that span a wider range of age groups. Schools exhibit a strong age structure with transmission pairs clustered along the main diagonal, whereas workplaces exhibit a high degree of homogeneous mixing across adult age groups, with reduced interactions involving younger individuals. Community settings primarily reflect the core peer diagonal with fewer secondary structures.

### Transmission Clusters

We study transmission clusters defined by shared membership of settings (household, family, school, workplace) defined by the nation-scale social network. We define such clusters by removing all edges in the network of plausible infections *G* for which the infector-infectee pair do not share one of the settings in the nation-scale social network. What remains is a set 𝒞 of connected components in which each component *c* ∈ 𝒞 represents a transmission cluster whose edges connect pairs of individuals belonging to the same setting. For example, a cluster could include a group of schoolchildren connected in *G* who are also connected to a household that one of these children belongs to, who are further connected to a workplace at which one parent works.

Across all major variants considered in our analysis, we identify 7151 such clusters, with cluster statistics shown in Figure 3. Panel A shows the spatial spread of the largest identified transmission cluster for each variant. Schools are the predominant setting of transmission in the largest clusters for the Alpha, Eta and Delta variants, whereas workplaces are the predominant setting for the largest B.1.177 and Omicron (BA.1) clusters. For the Omicron, Delta and B.1.177 variants, the largest clusters are located predominantly on the island of Zealand, where the capital Copenhagen is located, whereas the largest Alpha cluster is located predominantly in the north of Jutland, around the city of Aalborg. The largest Eta cluster is linked to a school outbreak in the city of Odense. Supplementary Figure 6 shows the temporal duration of these clusters for each variant, and whether the primary setting associated with each cluster was schools, workplaces, a combination of the two or “Other” (families and large households).

**Figure 3.**
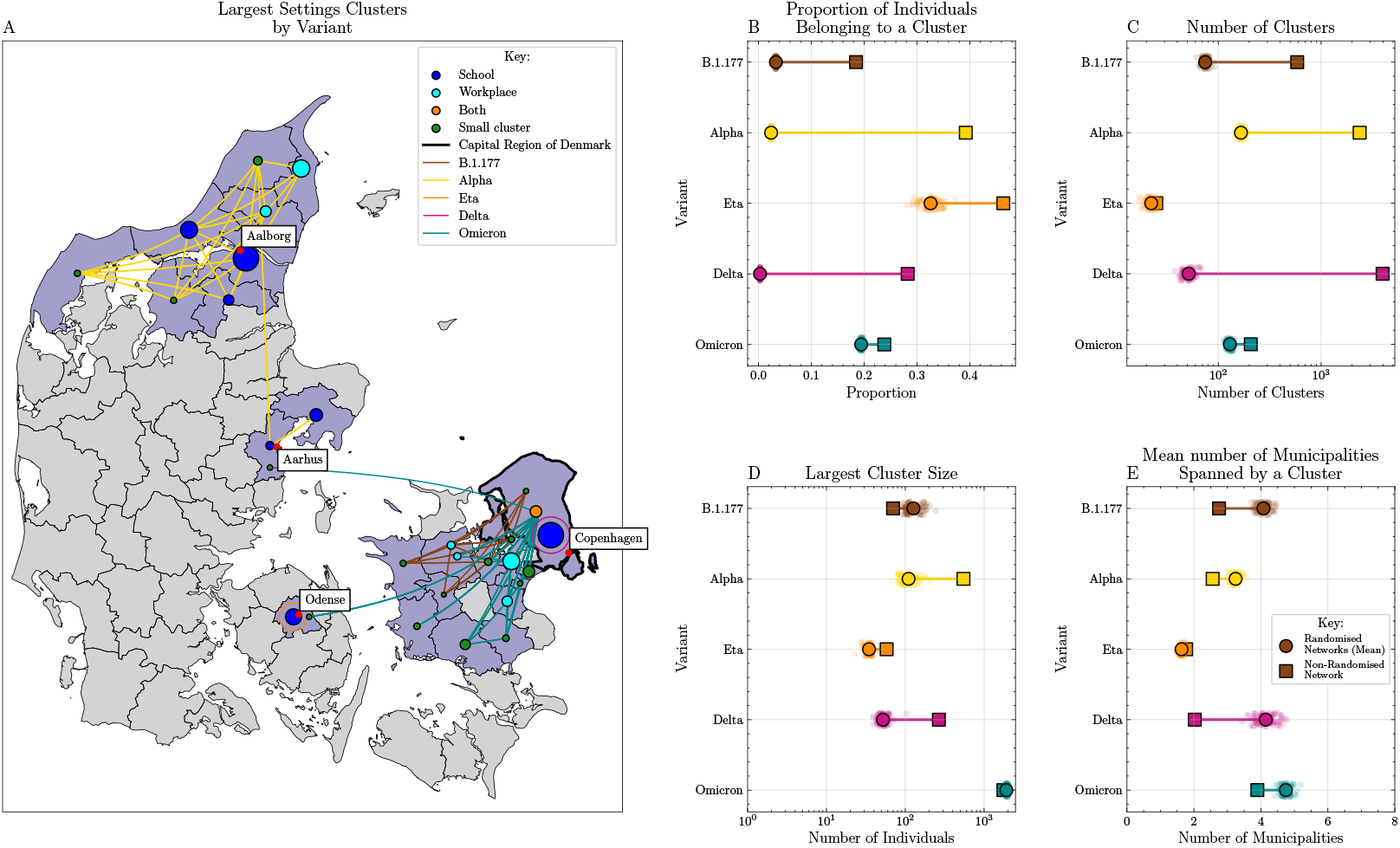
Transmission clusters: Largest settings-based cluster for each major variant in 2021. (A) Geographic spread of the largest cluster within each variant, with municipalities that have fewer than ten identified cases belonging to this cluster not displayed. Edges connect municipalities with five or more transmission pairs between them with edge colours representing the major variant to which the cluster belongs. Node colours represent the type of cluster (see definition of cluster types in Methods) and node sizes are proportional to the number of sequences within the cluster in each municipality. (B-E) Comparison of clusters derived from randomised networks with those from the inferred network of plausible transmissions. Squares represent the values obtained from the inferred network of plausible transmissions, with circles showing the mean values from 100 randomised networks, and with values from each randomised network scattered.

Figures 3B-E, show summary statistics for the clusters identified for each variant. To investigate the effect of including the genetic information in identifying these clusters, we compare these summary statistics with those from networks generated by randomly reallocating edges in *G* in a manner that ignores genetic similarity but preserves temporal consistency between plausible infectee-infector pairs. With the exception of Eta for some statistics, the summary statistics of clusters obtained directly from *G* differ significantly from those generated randomly, demonstrating that including genetic data in our analysis has a significant effect on our results. In particular, combining information on settings with genetic data results in identifying clusters that have a narrower geographic spread than those that do not make use of genetic data.

Combining genomic data with setting-based social network structure reveals strong variant-specific patterns in transmission environments, geographic spread, and cluster topology. Our analysis of Danish epidemic dynamics shows that transmission clusters are substantially more localized and structured than would be expected from temporal linkage alone.

### Case Reproduction Numbers and Overdispersion

Reproduction number estimates based on aggregate national reported cases and deaths [5, 23, 24] played a critical role in informing policy during the SARS-CoV-2 pandemic [25, 26]. From the reconstructed transmission networks, we can estimate the case reproduction number from the infections caused by each individual [27]. We then compare our estimates of the case reproduction number from individual data with those derived from national-aggregated data [28]; in principle, these two measures should align, and we show that this holds empirically during periods when large numbers of infections are sequenced.

During high sequencing periods, defined as periods when the sequencing proportion exceeds 30%, our adjusted instantaneous reproduction numbers, based on a renewal equation model using aggregate national data (Supplementary Information Section 4.1), closely match our case reproduction numbers, Figure 4A, with a correlation of 58% and a average of difference of 0.14 during the high sequencing period versus an average difference of 0.34 during the low sequencing period. During low-sequencing periods, defined as those with a sequencing proportion below 10%, we observe significant differences between the two estimates of the effective reproduction number. Early in the study period in 2020, estimates were based on small sample sizes and during a period of low case numbers, resulting in high variability in reproduction number estimates. The case reproduction number, based on individual-level data, is preferred because it better accounts for heterogeneity in transmission. We note that at the beginning of 2021, an estimate of *R*_*t*_ closer to 1 was a more realistic reflection of the pandemic at that time than an estimate of *R*_*t*_ *<* 0.5, which would have suggested an epidemic in rapid decline.

**Figure 4.**
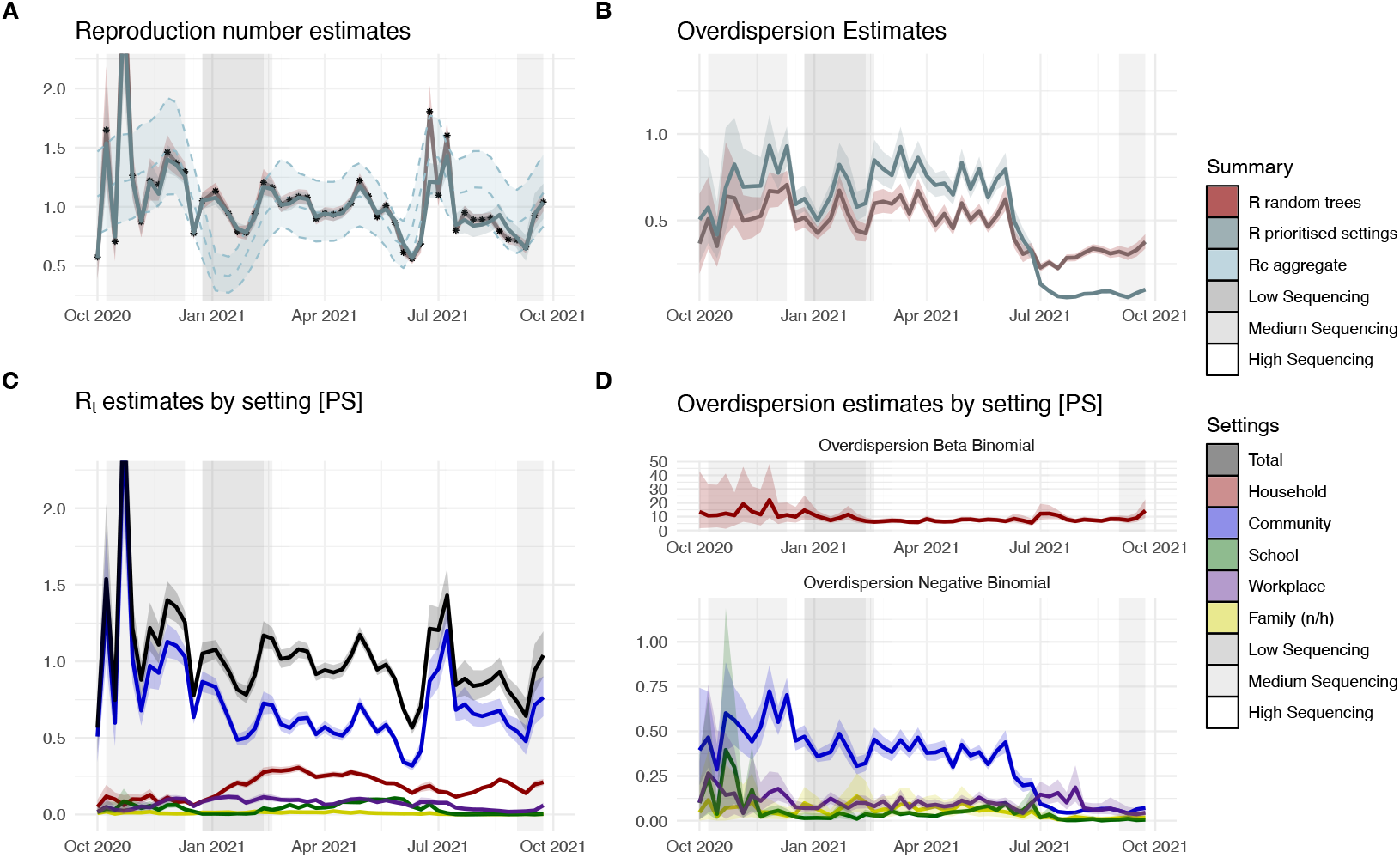
Effective reproduction numbers and overdispersion: (A) Comparison of aggregate case reproduction number estimates with estimates derived from random trees and prioritised trees, (B) Comparison of overdispersion estimates based on random and prioritised trees, (C) decomposition of prioritised settings case reproduction number by setting, and (D) overdispersion estimate for prioritised setting for households (beta binomial model) and schools, workplaces and the community (negative binomial model). Family (n/h) = Family not living in the same household as the infector.

We also decompose the reproduction number by setting (Figure 4C). In 2020, sequencing proportions are generally low, and we are unable to assign settings for most infector/infectee pairs; however, during the high-sequence period of 2021, we observe that household transmission accounts for approximately a third of overall transmissions [29, 30]. Workplaces and schools also each contribute over 10% of transmission events, albeit at a lower rate than suggested in the literature [29]. Family outside the household contributes very little to overall transmission, which may be due to the definition of family outside the household, which captures most core family transmission events already captured by household transmissions. The remaining transmission events are classified as *community* transmissions, which consistently account for the largest proportion of our *R*_*t*_ estimate.

The number of infectees per infector varied strongly, suggesting overdispersion or occasional superspreading. We characterise the offspring distribution using the secondary infections caused by an infector as *p*(*Z* = *n*), where *Z* 𝒟 ∼ (*v*) [31] and *v* is a random variable representing the individual reproduction number. The data show heterogeneity in *v* (Supplementary Figure 14), suggesting that the negative binomial distribution is best suited to model *Z*, with mean *R*_*t*_ and overdispersion parameter *k*. For the setting-specific analysis, we make a further modelling choice by using the beta-binomial distribution to model households, given that household sizes are finite and known. This allowed us to model the overdispersion parameter of the offspring distribution, and we observed that overdispersion increases (*k* decreases) with the arrival of the Delta variant (Figures 4B and 4C).

### Effectiveness of non-pharmaceutical restrictions based on individual level data

We use the inferred transmission networks to investigate setting-specific variation in the effectiveness of NPIs, as given by the Oxford Covid Government Response Tracker [32, 33] (see 4 for specifications of each model, as well as Supplementary Figure 13 and Tables 2-4). The specifications of each model are described in the Methods section.

By regressing individual-level reproduction numbers on the presence/absence or level of different interventions (non-pharmaceutical and vaccination status), we derive estimates of their effects on transmission. Figure 5 summarises our estimates for the prioritised settings trees (see Supplementary Figure 18 for results for the random trees). We compare results for the whole pandemic with those from the model run on sub-periods defined by sequencing proportions > 30% and near-maximal NPIs, that is, NPIs at either the maximal level or one level below the maximum restriction. In Supplementary Information Section 5.3, we present extensive sensitivity analyses examining prioritised versus random transmission networks, the impact of including the family setting, different levels of sequencing required, and a comparison of the maximum likelihood tree versus trees with random selections.

**Figure 5.**
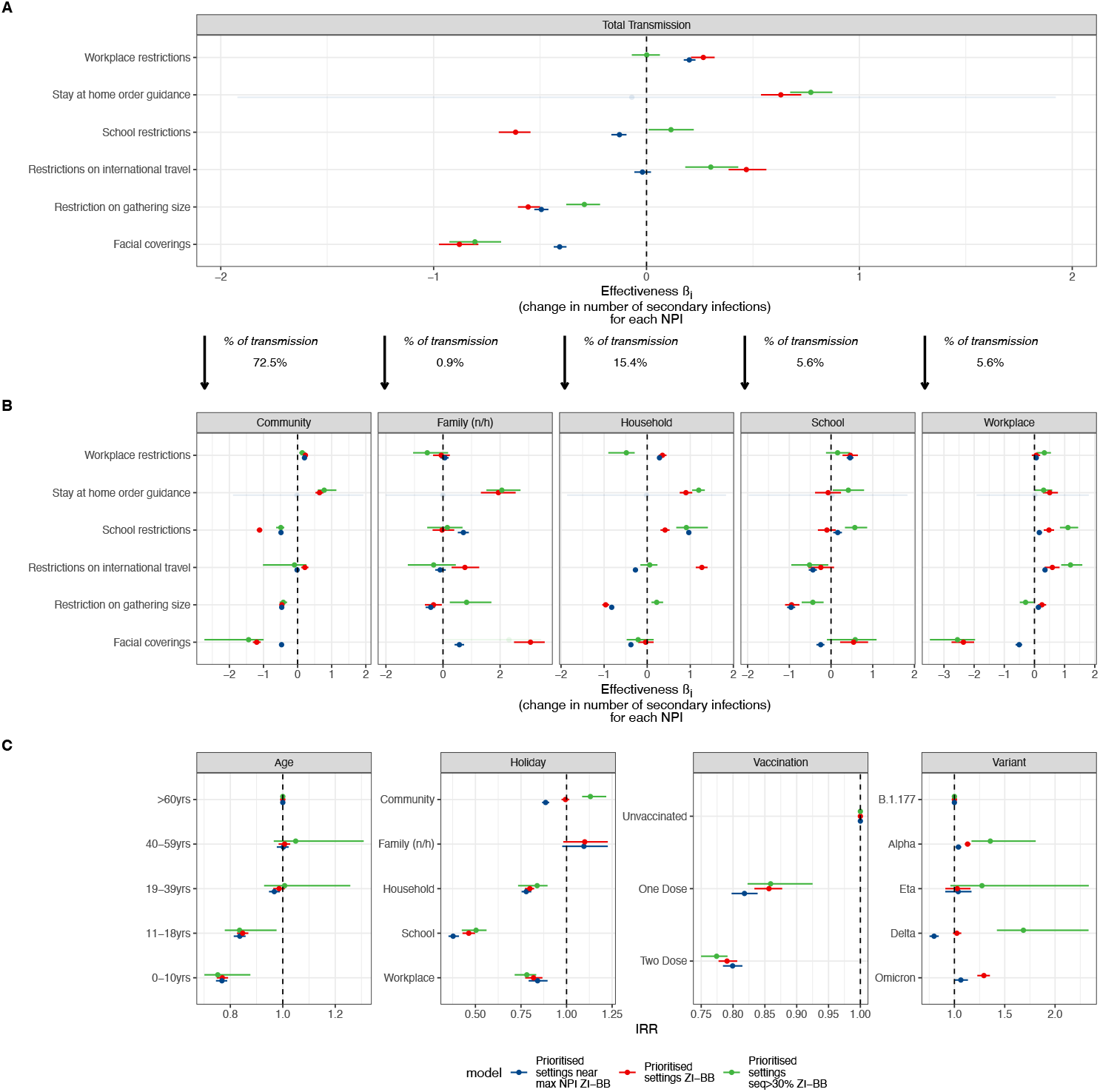
Effectiveness of NPIs based on individual-level data: (A) Effect sizes for the association between the number of infections caused by an infector and the level of each NPI across all transmissions. (B) Effect sizes for the association between the number of infections caused by an infector and the level of each NPI by setting. (C) Risk heterogeneities of the number of secondary infections by age, school holidays, vaccination status and SARS-CoV-2 variant. For all panels, the colours of the estimate correspond to the model displayed. Family (n/h) = Family not living in the same household as the infector. ZI-BB = Zero-Inflated Beta Binomial model for households only; a zero-inflated negative binomial model is fitted for other settings. The section between panel (A) and (B) indicates how many of the transmission events took place in that setting. The full results are available in Supplementary Table 5.

The impact of workplace restrictions, stay-at-home guidance, and restrictions on international travel was associated with increased overall transmission. In contrast, school restrictions, restrictions on gathering size, and restrictions on facial coverings were associated with reduced overall transmission (Figure 5A). The total transmission results are driven by the Community setting, as 72.5% of transmissions occurred there over the study period.

It is important to note that we only know that individuals are linked by a setting from the nation-scale social network, which is a static snapshot of social connections. This link does not imply that a transmission event necessarily took place in that setting. For example, two school-aged children may be an infector/infectee pair, but transmission could have occurred at home after school, during the weekend or during school holidays. School restrictions cover a range of measures (Supplementary Figures 11 and 12), particularly a phased return to in-person schooling [34] and automated circuit breakers [35] in schools, which had significant impacts on schooling [36]. For the default model, school restrictions were associated with a significant reduction in community transmission (−1.1, 95% highest posterior density interval [HDI]: −1.2 to −1.0) but an increase in household (0.4, 95% HDI: 0.3 to 0.5) and workplace (0.5, 95% HDI: 0.3 to 0.7) transmission. No significant changes in transmission were observed in schools or non-household family settings under the default model. The results for school restrictions are robust across time periods and settings, except that we observe an association with increased transmission in the school setting during the high sequencing period, consistent with the fact that most restrictions were removed during the second half of 2021 (Supplementary Figure 12). Similarly, we observed an association between workplace restrictions and a slight increase in transmission across all settings, except the family (non-household) setting, suggesting that workplace closures had little impact on transmission during a period of the pandemic when stay-at-home orders were not in place.

For facial coverings, we observed a significant reduction in transmission, regardless of model, in community settings (−1.2, 95% HDI: −1.3 to −1.1) and workplaces (−2.4, 95% HDI: −2.7 to −2.0). There was no significant impact on the household setting (0, 95% HDI: −0.2 to 0.1) and a significant association with increased family (non-household) transmission (3.1, 94% HDI: 2.5 to 3.6) and a much smaller effect in the schools setting (0.5, 95% HDI: 0.2 to 0.9).

Cancellation of public events was excluded from the analysis, as such measures were only present at the beginning of the study period. We did include stay-at-home orders, but note that these were only present at very low levels, and no strict ‘lockdown’ type measure was in place during the study period (Supplementary Figure 13), similar to most other European countries. The most relevant intervention was restrictions on gatherings, which were associated with a reduction in transmission in the community setting (−0.5, 95% HDI: −0.5 to −0.4), the family (non-household) setting (−0.3, 95% HDI: −0.6 to 0), the household setting (−1.0, 95% HDI: −1.0 to −0.9) and the schools setting (−0.9, 95% HDI: −1.1 to −0.8). Conversely, in the workplace setting, we observed a small association with increased transmission (0.2, 95% HDI: 0.1 to 0.4). International travel restrictions were associated with small changes in community transmission (0.2, 95% HDI: 0.1 to 0.3), households (1.3, 95% HDI: 1.1 to 1.4), and workplace settings (0.6, 95% HDI: 0.03 to 0.8). This suggests that these restrictions led to more movement and/or holidays within Denmark, keeping transmission local.

We observed that most NPIs (particularly stay-at-home orders, school restrictions, and gathering restrictions) were associated with increased family (non-household) transmission. It is important to note that the family (non-household) setting contributed minimally to overall transmission (less than 0.48% of all transmissions and less than 1.76% of transmissions within settings) and primarily affected individuals aged 20-59 years, suggesting that socialising among non-high-risk individuals may drive these transmissions (Figure 2A and F).

School holidays (Figure 5C, Holiday panel) had a significant impact on reducing transmission between individuals with a school link (IRR −53.6%, 95% HDI: −57.1 to −50.2) and on workplace-related reductions (IRR −18.1%, 95% HDI: −22.7 to −13.2). We also observed a decrease in the household setting (IRR −20.1%, 95% HDI: −22.8 to −17.8), potentially due to reduced outside household contact during holidays. Interestingly, an increase in the family (non-household) setting may have occurred during holiday visits, but we observed no significant results.

Our estimates of age-related variation in transmission risk (Figure 5C, Age panel) indicated that children and adolescents had less risk of transmitting onwards, relative to the 60+ reference age group, and that the 19-59 year age group had similar transmission risk, consistent with the findings in Goldstein *et al*. [37]. We provide plots of age-specific school- and work-testing rates, test positivity rates, and sampling proportions by group in the Supplementary Figure 2, which do not indicate bias due to testing variation.

For our default model, we found that vaccination reduced the risk of transmission by 15.5% (95% HDI: −18.2 to −13.1) after one dose and by 23.5% (95% HDI: −21.5 to −25.2) after two doses relative to the unvaccinated group as the reference, addressing a substantial gap in the literature highlighted by Mostaghimi *et al*. [38]. The median time since the second dose for our study population was 133 days, with an interquartile range of 102 to 167 days.

We did not observe significant regional random effects (Supplementary Figure 16); however, the pattern of the estimates was consistent with other research [39]. The one regional effect that was significant was higher community transmission in Hovedstaden (Capital Region of Denmark), consistent with the higher population density; the absolute number of infections was also higher in this region.

Lastly, we estimated overdispersion in the offspring distribution by setting (Supplementary Figure 16). We observed no overdispersion in the household setting, moderate overdispersion in the family (non-household) setting, and higher levels of overdispersion in the community, school and workplace settings, consistent with previous studies [40–42].

## Discussion

We used genetic distances between sequence pairs and testing dates to infer a network of plausible transmission pairs, and combined this network with data from the nation-scale social network to link plausible transmissions to settings such as households, workplaces, schools and family relations outside of the household. The resulting network showed temporal changes in transmission attributable to these settings as well as geographic clusters of transmission within settings showing evidence of localised spread. The age-structure that we find in contact patterns carry important implications for intervention strategies. For example, if the goal is to reduce broad cross-age mixing, measures targeted at workplaces may be particularly effective.

Since Bayesian phylogenetic reconstruction is infeasible at national scale with several hundred thousand sequences, we developed the methods used in our analysis to be computationally efficient. Furthermore, we found that relying solely on genetic and testing data produces transmission trees that substantially underestimate the proportion of transmission occurring in identifiable settings. We therefore recommend incorporating detailed contact information to improve inference of transmission pairs. Even when extensive registry and nation-scale network data are included, more than half of inferred transmission events remain unattributed to any known setting. This may exceed the true level of transmission occurring in the general community, although previous evidence suggests that casual community contacts played a meaningful role in the spread [16, 43].

We then inferred case reproduction numbers from the total number of secondary infections caused by each infector and found they were consistent with estimates derived from aggregated case counts, both our own model and models given in [23] (Supplementary Figure 10). The level of overdispersion we estimate, both from fitting the offspring distributions and the model estimating the effectiveness of NPIs, is consistent with the wider literature [40–42]. One advantage of our data is that we can estimate how overdispersion evolved over time, and we observed an increase in overdispersion as the Delta variant started to dominate in Denmark.

Our individual-based approach allows us to assess the impact of interventions on transmission, finding, for example, that two-dose vaccination, versus the reference group of unvaccinated individuals, reduced onward transmission by 23.5%. Previous studies, based on cohorts in the UK [44] and Israel [45], were at a much smaller scale and potentially suffered from under-ascertainment due to limitations in contact tracing. Past estimates from Israel are comparable to ours, with a 23% reduction in transmission risk, but have very wide confidence intervals [45]. Similarly, a UK modelling study using aggregate data found a decrease in transmission risk of 48.7% (95% CrI 28.0% - 71.3%) associated with vaccination after three doses but negligible effects after two doses [46].

Our results on the association between individual NPIs and total transmission (Figure 4A) need to be considered in the context that the study period was characterised by lower levels of restrictions than those in early 2020 at the start of the pandemic, and coincided with the rollout of vaccination. We found positive associations with total transmission for workplace restrictions, stay-at-home guidance, and international travel restrictions, which contrast with studies from the early period of the pandemic [7–9]. However, these restrictions were also predominantly guidance-based in Denmark, with no lockdown-style measures in place during our study period. Our findings that school restrictions, gathering-size restrictions, and facial coverings were associated with reductions in overall transmission align with the literature and are consistent with several systematic reviews on NPIs [47, 48].

Crucially, our approach also allows us to evaluate the effectiveness of NPIs on transmission across different settings, such as reductions in community and workplace transmission when facial covering mandates were in effect, or reductions in household and workplace transmission when large gatherings were restricted. School restrictions were negatively associated with community transmission but showed no association in the school setting. This result is consistent with the set of restrictions in place in schools during 2021 (Supplementary Information Section 4.2, [34, 49]), which were focused on reduced attendance in the first half of 2021 and hygiene measures in the second half of 2021. These measures may have had limited impact on in-school spread, but reduced attendance likely decreased mixing outside school, thereby lowering community transmission. Similarly, we observed that facial coverings were negatively associated with workplace and community transmissions but not in the family (non-household) or household setting. For schools, the results on facial coverings align with the lack of explicit rules on facemasks [34]. National scale studies that stratify the effects of NPIs are rare because they require population-wide data of the type used in this analysis. As a result, there is a paucity of comparable findings, and this remains an important area for future research.

One limitation of our study is missing data from individuals who tested positive that were never sequenced from infected individuals who were never tested, who thus are not represented in our network, *G*. Our estimates of the infection ascertainment rate remain high over the study period, particularly through 2021, and the same is true for sequencing rates, with some small deviations across age groups. Nevertheless, these individuals who are missing from our study may provide links between clusters that we have not been able to observe, thereby improving our ability to map the spread of SARS-CoV-2 between settings and to estimate community transmission.

A further limitation is that for some periods, due to the fast evolution of SARS-CoV-2, some individuals have a large number of plausible infectors, leading to misclassification error in determining the true infector. This would create substantial bias if we relied on randomly choosing infectors, and is mitigated by our choice to preferentially choose infectors from the same setting. Further misclassification could occur in the way that information on settings is recorded in our data. For example, workplace information was collected on the 30^th^ of November each year. Some people may have left the workplace between this date and the time that they were infected, whilst others may have joined, introducing spurious workplace edges whilst failing to capture others. Our estimates of NPI effectiveness are based on associations rather than a causal analysis and therefore do not constitute policy recommendations; instead, they provide information to inform subsequent work on potential policy options. More targeted analyses on individual interventions are better positioned to provide causal insights, for example, as in Espenhain et al. [35].

Overall, studying transmission in clusters in addition to sampling transmission trees offers a potential remedy to some of these limitations. Without deciding who infected whom, we can nevertheless study disease spread in settings and the geographic spread of transmission clusters, avoiding the aforementioned misclassification errors. This offers an approach to phylodynamic inference that can be readily applied to other pathogens and in other settings. Our hope is that this study can form a basis for establishing surveillance strategies for future infectious disease outbreaks, combining sequencing data with data on social contacts (including contact tracing) and other individual metadata. Our methods are fast, scalable, and could provide a cost-effective way to combine different sources of data collected in future pandemics to construct plausible routes of transmission and understand the effectiveness of NPIs as an outbreak develops.

## Methods

### Genomic and Testing Data

Denmark adopted a two-track testing strategy during the COVID-19 pandemic, consisting of a healthcare track and a community track. The healthcare track focused on testing SARS-CoV-2 amongst in- and out-patients (including regular screening of healthcare staff), whereas the community track consisted of free testing available on demand from members of the general public. The community testing track contributed to over 80% of tests conducted throughout the pandemic [50]. To facilitate this testing, Statens Serum Institut (SSI), the governmental public health institute in Denmark responsible for infectious disease control and prevention, established a large-scale testing laboratory (TestCenter Danmark) to analyse the results of PCR tests taken as part of the community track. Community testing centers were established such that every person had a testing centre within 20 km of their home address, and where PCR tests were performed by trained healthcare professionals. For further details, see [50]. Starting in February 2021, rapid antigen tests were also included as part of the community track for testing; however, a confirmatory PCR was required upon receipt of a positive result in order to be identified as a case for the purpose of national case numbers [50]. Further details on the sequencing protocols are given in the Supplementary Information Section 1.

### Registry Linkage

Pseudo-anonymised Danish civil registration numbers were used to link genetic sequences with individual metadata from the Danish registries, stored at Statistics Denmark. We used registry information on individuals’ age, sex, test date, vaccination status and registered home address. We also included the locations corresponding to each individual’s registered home address, in terms of NUTS 2 region (EU Nomenclature of Territorial Units for Statistics, namely: Hovedstaden, Midtjylland, Nordjylland, Syddanmark and Sjælland) and Danish municipality (*n* = 98).

### Danish Nation-Scale Social Network

The Danish Nation-Scale Social Network [51] is a population-level network that contains information on the relationships between individuals in the Danish registries. The network uses registry data collected each year in Denmark to identify an individual’s membership of the following social foci, i.e. entities around with joint activities are organized: households (based on address), schools, workplaces, families and neighbourhoods. Each household, school and workplace in the Danish registries is given a unique identifier, and the network of individuals belonging to these settings is represented in a bipartite manner. For a more detailed account of the construction of the nation-scale network, we refer readers to [51].

#### Households

We define household membership differently from the definition used in the nation-scale network to achieve greater temporal granularity. The original nation-scale network identifies an address as a household if there are either one or two individuals living at that address on the first of January of a given year, alongside any children that live in the household. Instead, we take a different, dynamic approach that defines a household based solely on membership of an address (regardless of the familial relationship between individuals living at that address) to account for shared housing between non-family members. This dynamic approach is preferable because it captures real changes in household composition throughout the year, rather than relying on a single annual snapshot, and it better represents non-traditional living arrangements such as shared housing among unrelated individuals. As a result, it more accurately reflects patterns of close contact that are relevant for transmission.

#### Schools

We construct a school network containing all information on individuals in primary and secondary education obtained from the Population Education Register. Individuals in this network are assigned a unique institution identifier for the educational institution they attend, along with a date marking any change of institution (for example, when moving from one school to another). We link individuals to their educational institution using a unique personal identifier.

#### Workplaces

We construct a workplace network from data collected from the Labour Market Register, indicating the workplace that each individual is registered as working at on the 30^th^ of November annually. For our analysis, which focuses primarily on the end of 2020 and the whole of 2021, we consider the workplaces that individuals were members of on the 30^th^ of November in 2020, again linking workplaces to individuals via a unique personal identifier.

#### Family Relationships

The nation-scale network also consists of information on whether two individuals have a family relationship. Family relationships are defined on the basis of child-parent relationships, and include the following relationships: child, grandchild, half sibling, full sibling, sibling (one parent shared, but information on other parent(s) unknown), cousin, co-parent, parent, grandparent, aunt/uncle and niece/nephew. For the purposes of our analysis, we consider only family relationships where the individuals do not also share a household, and consider those that do share a household to be household edges rather than family ones. We note that this choice may overlook additional family-based contact among related individuals who also co-reside. However, we prioritise household edges in these cases because shared residence represents a stronger and more sustained context for transmission than kinship alone, and this prevents double-counting of contact pathways.

#### Construction of a Subset of the Nation-Scale Network for 2021

We construct a subset of the full nation-scale network, which we will denote by 𝒩, that contains the information on social relationships that is relevant for our study. We denote the set of edges of by *E*(𝒩). For the purposes of constructing a network of plausible transmission pairs, we consider the household that an individual belongs to as being the address that they are registered at at the time of their first positive test. This amounts to drawing an edge between two individuals, (*v*_*i*_, *v*_*j*_) ∈ *E*(𝒩), if, and only if, they live at the same address at the time that *v*_*i*_ tests positive. Similarly for the schools network, we include the edge (*v*_*i*_, *v*_*j*_) in 𝒩 if *v*_*i*_ and *v*_*j*_ attend the same educational institution on the date that *v*_*i*_ first tests positive.

In the case of workplaces, we do not have sufficient temporal resolution to determine whether two individuals are in the same workplace at the time when one of them tests positive, so instead we include an edge (*v*_*i*_, *v*_*j*_) if *v*_*i*_ and *v*_*j*_ belonged to the same workplace on the 30^th^ of November 2020. This likely results in the misclassification of some pairs of individuals as colleagues. In our data, we find that 68.7% of individuals with a registered workplace on the 30^th^ of November in both 2020 and 2021 are registered as having the same workplace in both years, with 31.3% having different registered workplaces. High levels of job mobility are typical for Denmark [52], though this could also have been exacerbated by the pandemic. Since we do not have information about where in the year they changed their workplace, we assign all individuals to the workplace at which they were registered in November 2020. Finally, we connect all pairs (*v*_*i*_, *v*_*j*_) that are defined as family members (outside of the household) in 2021. This defines the subset, 𝒩, of the nation-scale social network that we use in our analysis.

### Between-Host Viral Evolutionary Model

We define the joint probability distribution of the observed Hamming distance between genetic sequences, *H*_*A,B*_, and the serial interval *T*_*A,B*_, conditional on an event where *A* infected *B*. We assume that, conditional on the transmission event *A* → *B*, viral evolution proceeds independently in the two hosts after infection. Furthermore, we assume that at the time of transmission the dominant viral variant in *B* is identical to the dominant variant in *A* at the moment at which *A* infects *B*.

We consider probabilities of the form:

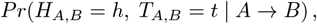

for both *h, t* ℤ _≥0_, under the assumptions that post-transmission mutations occur independently in *A* and *B*, and that the transmitted viral variant is the same as the variant corresponding to the dominant variant in *A* at the moment of transmission (effectively assuming a bottleneck size of 1). This is consistent with the assumption of a tight transmission bottleneck that has been confirmed for SARS-CoV-2 transmission by numerous studies [53, 54].

Let *T*_*A*_, *T*_*B*_ be the times at which the positive PCR test from which the genetic samples belonging to *A* and *B*, respectively, are performed, and let *H*_*A,B*_ be the Hamming distance between these genetic samples. We model the accumulation of mutations, which we refer to as substitutions, that occurs in the time between *T*_*A*_ and *T*_*B*_ as a homogeneous Poisson process, for which we assume a substitution rate of *µ* = 0.091 substitutions per day (equivalent to a rate of 3.04 × 10^−6^ substitutions per site per day) such that, on average, a single substitution is accrued after 11 days. This closely matches the substitution rate of 1.39 × 10^−3^ substitutions per site per year identified in [17]. We further assume that the random time interval, *I*, between an individual being infected and taking their first positive test follows a lognormal distribution:

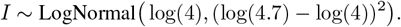

This parameterisation is taken from a meta-analysis of estimates of the incubation period for SARS-CoV-2 [18, 55]. In practice, since we measure time discretely, we use a discretisation of the lognormal distribution, defining the discrete distribution function *p*_*I*_ (*x*)= *Pr*(*x* - 0.5 *< I < x* + 0.5).

*H*_*A,B*_ depends on whether *B* was infected before or after *A* tests positive or not. If *B* was infected before *A* tests positive, then viral evolution occurs in both individuals until *A* tests positive, after which point we only observe the evolution that occurs in person *B*. If *B* was infected after *A* tests positive, then we only observe the evolution that results from viral evolution that occurs in *A* until the moment of transmission, after which the evolution continues in *B* until the moment that *B*’s viral sample is taken. In the second scenario, the average number of substitutions is *µ*(*T*_*B*_ - *T*_*A*_), whilst in the first it is *µ*(2*I* - (*T*_*B*_ - *T*_*A*_)). The number of substitutions *H*_*A,B*_ for a transmission pair (*A, B*) is therefore a linear combination of Poisson random variables, summing over all possible times at which *B* was infected relative to the time at which *A* was sampled:

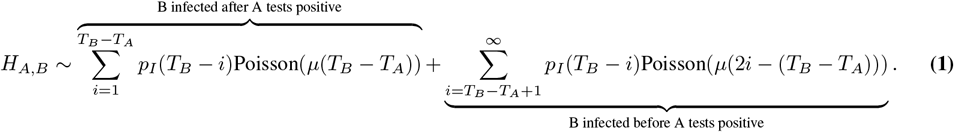

This gives us one component of the joint probability distribution for testing whether transmission between *A* and *B* is plausible, given the data. The other component comes from using *D*_*A*_ and *D*_*B*_, the dates at which the infection episodes of *A* and *B*, respectively, begin. Often, we have that *D*_*A*_ = *T*_*A*_ and *D*_*B*_ = *T*_*B*_, since the episode start coincides with the first positive PCR test taken by the individual during their infection, which is then sequenced. However, in some cases, an infected person takes a positive antigen test prior to having their genetic sample taken, or else the PCR test corresponding to the genetic sample for an individual does not come from the first positive test for the episode, in which case *D*_*A*_ and *D*_*B*_ are taken to be the earliest possible dates for each individual’s episode. We assume that the time between the first positive tests of an infector and their infectee follows the same distribution as the generation time interval (that is, the time between the infection of an infector and their infectee). We therefore assume that *D*_*A,B*_:= *D*_*B*_ - *D*_*A*_ for each pair are observations of a random variable *D*, which follows a Gamma distribution [18], so that *D* ∼ Gamma(4.87, 1.98), where the parameters of the Gamma distribution represent the mean and variance. Given these two models, we have that the probability of observing the data *D*_*A,B*_, *T*_*B*_ −*T*_*A*_ and *H*_*A,B*_ for a transmission pair (*A, B*) is given by:

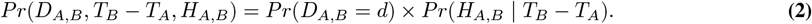

### Identification of Network of Plausible Transmission Pairs

Our aim is to construct a network whose nodes are drawn from the population *V* = {*v*_1_,... *v*_*N*_}, where individuals are connected if there is a plausible transmission link between them. We therefore identify a pair of individuals, (*A, B*) ∈ *V* × *V* as a *plausible transmission pair* whenever their genetic data and testing data are consistent with transmission having occurred from one individual to another, according to our model outlined in the previous section. Under this model, we calculate for each pair of individuals the probability, assuming that transmission has occurred, of observing the data associated with each pair. We then define a threshold probability (1 - *α*) such that we rule out a transmission pair as *implausible* if *Pr*(*D*_*A,B*_ *> d*, (*T*_*B*_ - *T*_*A*_) *> δt, H*_*A,B*_ *> h*) *< α*, and take *α* = 0.05 in our analysis. Conversely, if *Pr*(*D*_*A,B*_ ≤ *d*, (*T*_*B*_ - *T*_*A*_) ≤ *δt, H*_*A,B*_ ≤ *h*) ≥ 0.95, we conclude that the data are consistent with transmission from *A* to *B*, and label the pair (*A, B*) as a *plausible* transmission pair.

The collection of all plausible transmission pairs in our dataset gives rise to a directed network *G* of plausible transmission pairs with node set *V* = {*v*_1_,..., *v*_*N*_} and edge set *E* = {*e*_1_,... *e*_*M*_}. To each of these edges (*v*_*i*_.*v*_*j*_) is assigned a weight 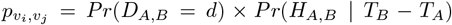, which is the probability under our model of observing the data given that the person represented by node *v*_*i*_ infected the person represented by *v*_*j*_. We further assign attributes to the nodes and edges corresponding to metadata relevant for our analysis. For each node, we take the time 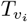 at which the test corresponding to the sequence represented by *v*_*i*_ was taken, and we attach as a node attribute unique identifiers for the settings (namely, household, workplace, school and family), if they exist, that the individual belongs to at time 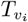. We also include as part of the node attributes the region and municipality to which the individual is registered as living in at time 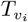. We then include as edge attributes for an edge (*v*_*i*_, *v*_*j*_) binary variables encoding whether the individuals represented by *v*_*i*_ and *v*_*j*_ share each of the four settings at time 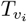. Edges between individuals ((*v*_*i*_, *v*_*j*_)) in this network therefore contain a weighting that is given by 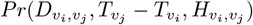, as well as any information on the relationship between *v*_*i*_ and *v*_*j*_ based on settings. A visualisation of the weighting between individuals identified as a plausible transmission pair is given in Supplementary Figure 5.

Note that the constructed network *G* is not a tree, and may even contain cycles if there is a non-zero probability of a transmission pair testing positive on the same day. Thus it should not be interpreted as a transmission tree, but rather a weighted network of plausible transmission pairs that contains information about potential transmission pathways.

### Sampling and Analysis of Transmission Trees

Below we proceed with strategies for sampling a single infector from the set of plausible infectors, 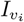 (represented as in-edges) belonging to each individual *v*_*i*_ in the network *G*. These strategies all depend on the probabilities 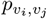 assigned as edge weights between individuals *v*_*i*_ and *v*_*j*_. Note that, for a given individual, the sum of the in-edge probability weights is not equal to 1 (i.e. 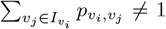). However, each of the probabilities has been calculated on an identical copy of the same underlying probability space, and so they may be compared with one another. The result of each of these sampling strategies is a collection of transmission trees in which each individual has either one infector or no infectors.

#### Maximum-Likelihood Infector Sampling

One sampling strategy, which we refer to as *maximum-likelihood infector sampling*, is as follows. For each individual *v*_*i*_, choose a plausible infector 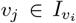 with the maximum edge weight i.e. such that 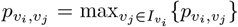. If this choice of *v*_*j*_ is not unique, i.e. if there are multiple candidate infectors that have the same maximum edge weight, then we choose uniformly at random from amongst those candidate infectors.

#### Random Infector Sampling

A second strategy, which we refer to as *random infector sampling* consists of choosing a plausible infector 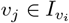 randomly with probability:

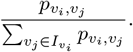

This ensures that candidate infectors are reweighted according to their edge weights and the number of potential infectors that *v*_*i*_ has. The resulting transmission trees sampled under this strategy are referred to as *random trees*.

#### Random Infector Sampling with Prioritised Within-Setting Transmission

We adapt the random infector sampling strategy described previously in order to choose potential infectors 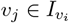 preferentially if they share a setting with *v*_*i*_ as defined in the nation-scale social network, that is, if (*v*_*i*_, *v*_*j*_) is also an edge in 𝒩. This amounts to restricting the set of plausible infectors for *v*_*i*_ to 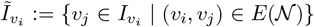 if 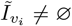, otherwise sampling randomly according to the edge weights (as done for random infector sampling) from the set of plausible infectors 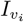 if there are no plausible infectors for *v*_*i*_ in any of the settings.

If 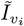 contains more than one plausible infector that shares a setting with *v*_*i*_, then we first choose as the sampled infector any of these infectors that shares a household (i.e., an address) with *v*_*i*_ (if one exists, choosing randomly between them according to their edge weights if multiple exist). We continue sampling preferentially from settings in the following order after households: schools, then workplaces, then families. If there is no plausible infector within any of the settings occupied by *v*_*i*_, then we sample an infector 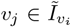 randomly according to the edge weights (*v*_*i*_, *v*_*j*_) from the set of all plausible infectors of *v*_*i*_. We refer to the collection of trees that are sampled via this method as *prioritised settings trees*, and it is these trees that form the main part of our analysis in Figure 2.

### Settings-based Transmission Clusters

The above approach to studying the network *G* requires us to choose a strategy for sampling a single infector from the range of plausible candidates that might be available for each individual, and is sensitive to that strategy.

As an alternative to having to choose such a strategy, we proceed by identifying transmission clusters from the network *G*, for which we are not required to sample an infector for each individual. We investigate how clusters of individuals with genetically similar viral samples interact with clusters identified in settings (households, schools, workplaces and family). In order to do this, we take take the intersection of the nation-scale network *N* with the plausible transmission network *G*. This is equivalent to removing edges between pairs of individuals that are not connected by any setting defined in 𝒩, and removing any nodes that do not have any outgoing or incoming edges as a result of this construction. The resulting graph *V*_*𝒩*_ is a subgraph of *G* in which all plausible infector-infectee pairs share an edge in 𝒩. We then take the collection of connected components of *V*_*N*_ that consist of greater than 5 individuals, *C*(*V*_*𝒩*_)= {*C*_1_, *C*_2_,... *C*_*m*_}, which we define to be *settings-based transmission clusters*.

We then investigate properties of the (settings-based) transmission clusters to inform how SARS-CoV-2 spread in Denmark according to the social-network structure given by 𝒩. Separating clusters by major variant (B.1.1.7, Alpha, Delta, Omicron and Eta), we calculate summary statistics of the resulting clusters. We firstly calculate the total number of individuals for each variant that belong to a transmission cluster of size greater than 5 as a proportion of the total number of sequences collected for that variant. We also report the total number of clusters (i.e. *m*), as well as the size of the largest cluster, identified for each variant. We also consider the geographic spread spanned by these clusters, both at the smaller municipality (total of 98 municipalities in Denmark) scale and the larger region scale (total of 5 regions in Denmark). We then calculate the average number of municipalities spanned by all clusters within a major variant classification, as well as the proportion of clusters that span more than one region.

We further classify clusters based on the predominant setting represented in each cluster. For instance, if the largest setting in a cluster is a school, and more than 5 individuals belong to the same school, then we identify the cluster as a school cluster. The choice of 5 individuals as a cutoff is taken to preserve anonymity of individuals identified in the transmission cluster. If, additionally, there are more than 20 individuals belonging to the same school and another group of more than 20 individuals belonging to the same workplace within a cluster, the the cluster is identified as both a school and workplace cluster. If fewer than 5 individuals belong to a single school or workplace, then the cluster is classified as a small settings cluster (consisting of only one or a small number of households or families connected by transmission in other settings).

To verify the signals observed from these clusters we also generate randomised clusters obtained by randomly reassigning edges of the plausible transmission network *V*. We fix the out-degree distribution of *V* and reassign outgoing edges by sampling uniformly at random from all available nodes representing individuals that received their first positive test within 11 days of the first positive test associated with the parent (infector) node. The choice of 11 days as a cutoff is to ensure that reassigned plausible transmission pairs are still within the 95^th^ percentile of our chosen generation time distribution. The effect of this is to remove the information on infector-infectee pairs that is inferred from the genetic data and to instead only assign pairs based on the timing between tests. For each generated randomised network *V* ^*i*^, we generate a set of clusters 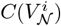 in the same way as for the true network and compare the summary statistics calculated for *C*(*V*_𝒩_) with the distribution of statistics calculated for this collection of randomised clusters.

### National Case Counts, Transmission Model and Infection Ascertainment Rates

Case counts and COVID-specific mortality numbers were sourced from weekly reports by the *Statens Serum Institut* (SSI). A semi-mechanistic branching process model, using daily case counts *C*_*t*_, which are modelled as *c*_*t*_ = 𝔼 [*C*_*t*_], and reported deaths *D*_*t*_ was used to model the disease transmission dynamics [24]. A component was added to the model to infer the time-varying weekly Incidence Ascertainment Ratios (*IAR*) following the approach of Mishra *et al*. [56]. In line with [56], we assume that the distribution *π*^*i*2*c*^, the time lag between infection and case identification, is zero for the first 3 days and 10% for the subsequent 10 days. The generation time distribution *g* was unknown but was approximated with the distribution of the serial interval, which was assumed to be Gamma distributed *g* ∼ Γ (6.5, 0.62) [8]. This allowed us to link observed cases with infections and infer the *IAR* (Equation 3). We defined the iar as a weekly random walk with a link function (Equation 4), specifically twice the sigmoid function *f* = 2 exp(*x*)*/*(1 + exp(*x*)).

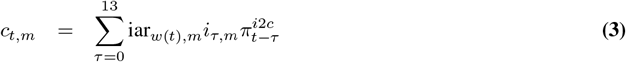

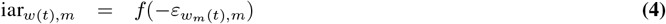

The parameters of the hierarchical model were jointly estimated, and inference was performed in R using Stan [57, 58] (version 2.32.5). The model is described in Supplementary Information Section 4.1.

### Comparison of effective numbers: instantaneous vs case reproduction numbers

We model the distribution of secondary infections caused by an individual using several distributions (Supplementary Figure 14), including Poisson, Negative Binomial, Zero-inflated Negative Binomial, and Negative Binomial distributions adjusted for partial observations. We fit these distributions to the inferred number of infections caused by an individual, as described in Sampling and Analysis of Transmission Trees. The case reproduction number 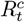 at time *t* is computed for the full time period (full data) and for weekly cohorts, that is, individuals who tested positive in that week (the number of infections caused by those individuals is not limited to that week but includes all infections).

To derive an estimate of *R*^*c*^ from the *R*^*i*^ estimated from aggregate case data, as described in section 4, we use the following transform, as defined in [28, 59]

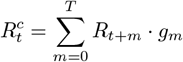

where *g* is the generation time distribution, as previously defined.

We fitted a range of distributions to the number of offspring infections per infector during a given week. The distributions included Poisson, Zero-inflated Poisson, Negative Binomial, and Zero-inflated Negative Binomial. The case reproduction number was taken to be the mean for each distribution. The Negative Binomial distributions also allowed us to quantify the offspring distribution’s overdispersion.

### Estimation of non-pharmaceutical intervention effectiveness from individual level data

In this section, we describe the regression model used to estimate the association between NPIs and the number of infections caused by an individual *y*, accounting for individual-level information. We consider models across four settings of interest: households, families outside the household, schools, workplaces, and community transmissions, which encompass any transmission event that cannot be attributed to another setting. The total number of infections caused by an individual is the sum of infections across settings (*y* = ∑_*s*_ *y*_*s*_), which we obtain from the transmission trees described above.

#### Data and Distributions

For each setting *s* ∈ {household, family (n/h), school, workplace, community}, where (*n/h*) is the abbreviation for non-household, we observe count data *y*_*s*_ of independent random variables *Y*_*s*_ with:

- Family (non-household), school, workplace, and community infections:

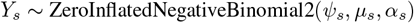

where *ψ*_*s*_ is the zero inflation probability for setting *s, µ*_*s*_ the mean of the negative binomial for setting *s* and *α* _*s*_ the overdispersion parameter. *µ*_*s*_ is modelled using linear predictors, see the subsection below.
- Household infections are modelled using a Beta-Binomial as the size of each household is known from the nation-scale network data, with a median of 3 household members (Supplementary Figure 17):

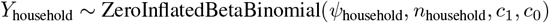

where *ψ*_household_ is the zero inflation probability and *n*_household_ the household size:

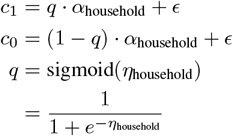

with *ϵ* = 10^−6^ for numerical stability and *η*_household_ is modelled as detailed in the linear predictors subsection. The parameters *c*_0_ and *c*_1_ are the shape parameters of the Beta-Binomial distribution that governs the number of household infections, conditional on the count not being zero-inflated. The expected household infection probability is given by 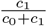, and the level of dispersion around this mean is given by *α*_*household*_ = *c*_0_ +*c*_1_, with large values of *α*_*household*_ implying low overdispersion and smaller values implying higher levels of overdispersion. *c*_0_ represents the concentration associated with expected non-infections in the household, and *c*_1_ represents the concentration associated with infections.

#### Zero-Inflation Component

For all settings:

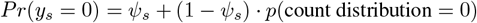

With a prior on the hurdle probabilities *ψ* for each setting:

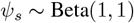

#### Linear Predictors

For each setting *s*:

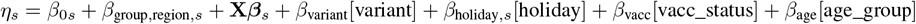

where for Negative Binomial outcomes 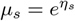.

In the model, we consider the following categorical covariates. We consider Wildtype, Alpha, Eta, Delta, Omicron for the variant periods, school holiday as either Yes or No for each date, vaccination status (*vacc*_*status*) includes unvaccinated, one dose, two doses, and we consider 0-10 yrs, 11-18 yrs, 19-39 yrs, 40-59 yrs, 60+ yrs as age groups.

The coefficients for the holiday covariate are allowed to vary by setting; however, the other covariates will have consistent estimates *across* all settings, as they are biological/epidemiological covariates which should be invariant to setting.

**X** is the design matrix for the covariates for NPIs (as defined in Tables 2 to 4):

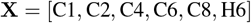

The NPIs are lagged by 5 days to account for the delay between infection and testing [7] and are scaled to the range 0-1.

We modelled regions as random effects to allow partial pooling across regions, thereby obtaining more stable estimates and reducing the risk of overfitting.

#### Priors

We used the following priors, in addition to previously stated priors in this section:

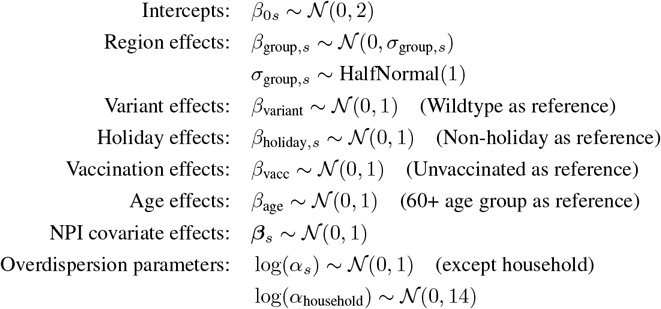

#### Notes

For household contacts, *n*_household_ is the household size. All categorical variables use reference coding in Python, and the model contains no interaction terms.

We excluded ‘cancellation of public events’ from our main analysis, which had no restriction except at the beginning of October 2020, and ‘protection of elderly people’, as that measure overlapped almost perfectly with the ‘facial coverings’ restriction. In the model, the restrictions were rescaled to the range of 0 to 1.

We estimate the model on the full study period. However, for sensitivity analysis, we also run the model on a selection of subperiods. These subperiods are defined by the proportion of infections that are sequenced (for example, sequencing proportion greater than 30% as shown in Figure 4); that is, we consider only days for which the sequencing proportion exceeds a given threshold. For these days, we include all secondary infections of an infector who tests positive on an included day, even if those infections take place on a non-included day.

Maximum NPIs are NPIs at the maximum level, as defined for each NPIs in Supplementary Tables D.2-D.4, with a value of one and zero otherwise, and near maximum NPIs are NPIs at 66% or more of the maximum level for each NPI, with a value of one and zero otherwise.

## Declarations

### Data availability

The data utilised in this study is accessible under restricted conditions under Danish data protection laws. Researchers can request access to the data from The Danish Health Data Authority, Statistics Denmark and Statens Serum Institut, complying with Danish data protection regulations and any necessary permissions. No data collection or sequencing was conducted specifically for this study.

### Code availability

All code relevant to reproducing the experiments is available online: https://github.com/JCurran-Sebastian/Danish_Transmission_NPIs_Cov19.

## Author contributions

J.C.S, C.M., J.J. and S.B. conceptualised the study. L.H.M., N.M.F. and S.B. supervised. J.C.S, C.M. and J.J. wrote software, and performed the analysis. J.C.S. and C.M. wrote the original draft and prepared all tables and figures. L.H.M, T.G.K., F.T.M., P.J., M.R., M.S., A.H. and J.F. were responsible for data curation and management. All authors contributed to critically reviewing and editing the manuscript.

## Ethical statement

This study is based on secondary analysis of administrative data collected by government agencies. The legal mandates to collect and assess the data are held by the agencies. The Danish Act on Ethics Review of Health Research Projects and Health Data Research Projects (“the Committee Act”) do not apply to the type of research reported in this paper. The study has been conducted in compliance with the General Data Protection Regulation of the European Union and the Danish Data Protection Act, which are enforced by the Danish Data Protection Agency. All data management and analyses were carried out on Statistics Denmark’s secure research servers. The study only contains aggregated results and no personal data.

## Acknowledgements

The authors express gratitude to Statens Serum Institut and The Danish Health Data Authority for their efforts in gathering and granting access to the data. Additionally, we are grateful to the Danish Covid-19 Genome Consortium (DCGC) for conducting the sequencing of SARS-CoV-2 samples; the full list of DCGC members and their affiliations is listed in the Supplementary Information.

