## Supplementary Information for "Transmission Networks and Intervention Effects From SARS-CoV-2 Genomic and Social Network Data in Denmark"

### Appendix 1: Ascertainment, Testing and Sequencing in Denmark

Here we present additional methods, results and figures relating to the levels of case ascertainment, rate of testing and proportion of positive RT-PCR tests that were selected for sequencing. Taken together, these results give a picture of the high level of surveillance in Denmark, particularly in 2021, which demonstrates that a large proportion of infected individuals are sequenced, and are therefore included in our study.

#### 1.1. Genomic Sequencing Protocols

Genomic sequencing was conducted by the Danish COVID-19 Geonme Consortium (DCGC). A full description of the protocols followed for the sequences in our data is given in [1], but we summarise some of the key details here. All sequences included in our dataset met the inclusion criteria set by the DCGC on the cycle threshold (Ct) value, which varied between 30 and 38 during the study period [1]. Genomic sequencing was conducted mostly by Aalborg University (AAU) with contributions from Statens Serum Institut (SSI) and regional clinical microbiology laboratories. Whole genome amplification of SARS-CoV-2 was conducted using a modified version of the ARTIC tiled PCR scheme, targeting 33 overlapping amplicons ranging between 1000 and 1500 base pairs [2]. A custom 2-step PCR strategie was used for barcoding the amplicon libraries, which then underwent normalisation, pooling and preparation for sequencing using the SQK-LSK109 ligation kit from Oxford Nonopore [3]. Sequencing was performed on the MinION device using R.9.4.1 flow cells from Oxford Nanopore. Base calling of the raw sequencing data was done using Guppy v.3.6.1 and demultiplexing was conducted using a custom cutadapt v.2.10 wrapper [4]. Consensus sequences were generated using the artic minion function, using default settings from the ARTIC network protocol (v.1.1.0), and consensus calling was done using medaka [5].

Further seqecning workflows included using the ARCTIC v3 amplicon sequencing panel, which consisted of 98 overlapping amplicons containing approximately 300 nucleotides [6]. In some cases, custom spike-ins were included to maintain consistent coverage [7]. This was done using Illumina platforms, such as the NextSeq or NovaSeq. Paired read lengths ranged from 51 to 150 nucleotides (wth 74 nucleotides being the mode). trim-galore v.0.6.10 was used to trim the sequencing reads, and consensus sequences were generated using a combination of iVar (v.1.4.3) and BCFtools v.1.18 commands [8, 9]. A PHRED score threshold of 20 was applied during read and primer trimming. The quality criteria required that sequences had fewer than 3000314 ambiguous bases (N's), $\leq 5$ ambiguous base calls, high yield relative to controls and that they passed quality control standards that detected contamination.

Sequences were aligned to the Wuhan WIV04 (MN996528.1) reference genome using MAFFT v.7.520. Problematic regions identified by de Maio et al. [10] were masked using the augur mask tool from the Nextstrain pipeline [11] (Augur version v.22.3.0), which included masking regions at positions 1-55 and from site 28804 onwards.

Infection episodes were defined by SSI as a 60-day window following the first positive PCR test for an individual. In our dataset we include a single genomic sequence per infection episode, corresponding to the highest quality sequence (i.e. with the lowest number of ambiguous sites) from within the episode. We then include in our dataset sequences corresponding to 5 different variants, namely: B.1.177, Alpha, Delta, Eta, and Omicron (BA.1) variants. In total, between September 1st 2020 and December 31st 2021, we include 293,841 SARS-CoV-2 genomes in our final dataset.

#### 1.2. Infection Ascertainment and Sequencing Proportion

We calculate the daily Infection Ascertainment Rate (IAR) as described in Section 4, the result of which can be seen in Figure 1. Between March and August of 2021, we find that the IAR is consistently above 60%, dropping somewhat after September 2021 before dropping to lower than 20% in December. This can largely be explained by the arrival of the Omicron variant, which resulted in a large wave of infections and, ultimately, in Denmark recording higher levels of incidence than had been seen in any previous wave of infections.

Similarly, Figure 1 shows that the proportion of positive RT-PCR tests that were sequenced over 2021 was high, consistently over 60% between mid-January and mid-November of 2021. The total proportion of positive RT-PCR tests that were subsequently sequenced over the study period is 48%. Combining the sequencing proportion with the IAR, we estimate the daily proportion of infections that were sequenced. This is represented by the blue curve in Figure 1, in which, for each day, we take the product of the sequencing proportion and IAR over the previous 11 days, which is the

95% percentile cutoff for the generation time distribution assumed in our baseline model of transmission. This gives us an estimate of the proportion of people infected within a generation time before each individual that are included in our data, and that can therefore be identified as an individual's plausible infectors.

Figure 2 shows testing and sequencing proportions broken down by different population groups. Firstly, we calculate the proportion of individuals in the population that take a test (either antigen or RT-PCR test) per day. Secondly, we calculate the proportion of those tests that are returned as positive. We finally calculate, as before, the proportion of positive RT-PCR tests that were sequenced. We then break down each of these proportions by different groups of the population, namely by age group, and then by whether or not the individual tested belongs to a school or a workplace, as defined by the nation-scale social network. Whilst our results show some between-group differences in the decision to take a test over time, they show a large degree of consistency in the proportion of tests that are positive and in the sequencing proportion. The exception to this is in the oldest (60+) and youngest ($\leq 10$) age groups, which show a lower sequencing proportion of positive RT-PCRs compared with other age groups.

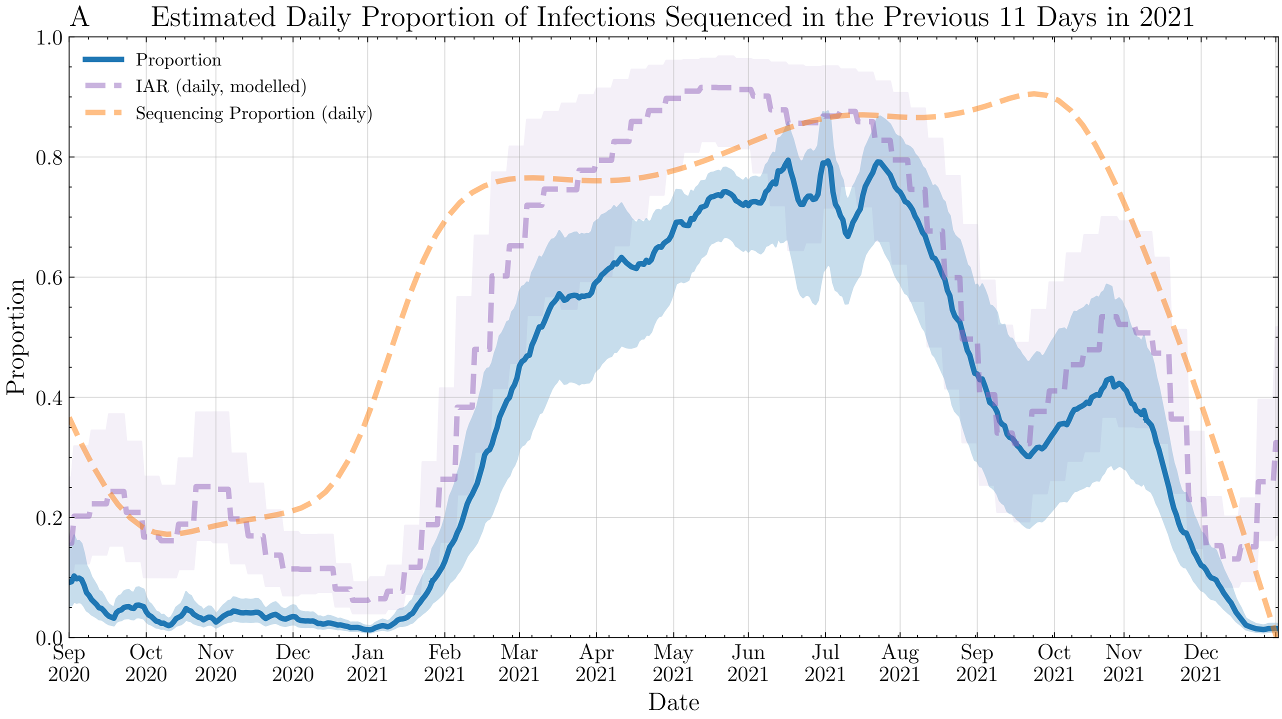


**Supplementary Figure 1.** Inferred Infection Ascertainment Ratio (IAR, purple) and daily proportion of positive PCRs that were sequenced (orange). The blue curve shows, for each day, the proportion of infections occurring within the previous 11 days that were identified as cases and then subsequently sequenced.

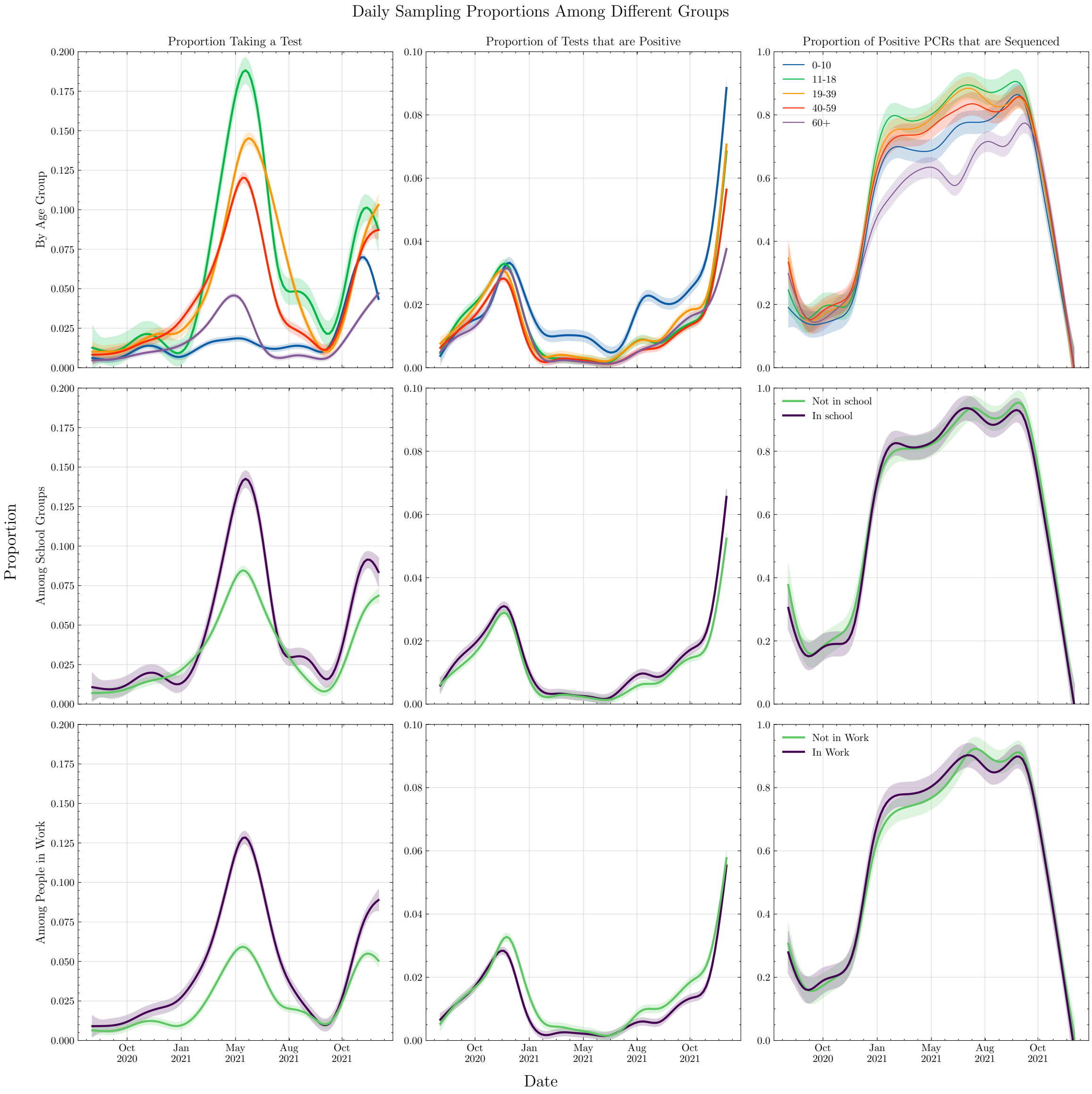


**Supplementary Figure 2.** *Left column:* Daily proportion of individuals in the population that take a test (RT-PCR or antigen). *Middle column:* Daily proportion of tests that are positive. *Right column:* Daily proportion of positive RT-PCR tests that are sequenced. Daily proportions are separated by age group (*top row*), education status (*middle row*) and employment status (*bottom row*).

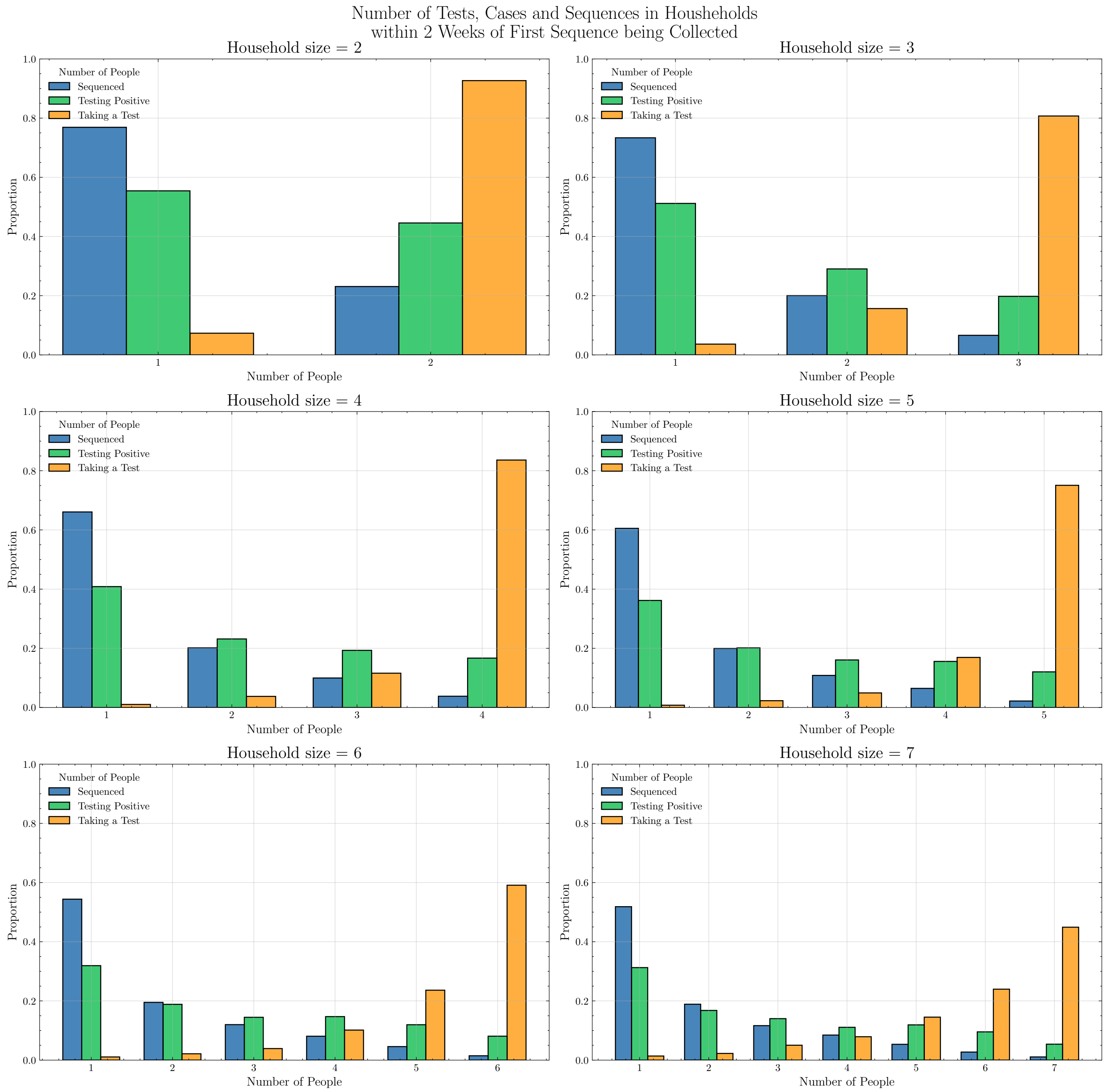


**Supplementary Figure 3.** Proportions of individuals in a household where an individual has been sequenced for SARS-CoV-2 that take a test (antigen or RT-PCR), test positive, and have a positive RT-PCR test sequenced within 14 days before or after the first sequence in a household outbreak is collected. Results separated by the size of the household at the time that the RT-PCR test giving rise to the first sequence is taken.

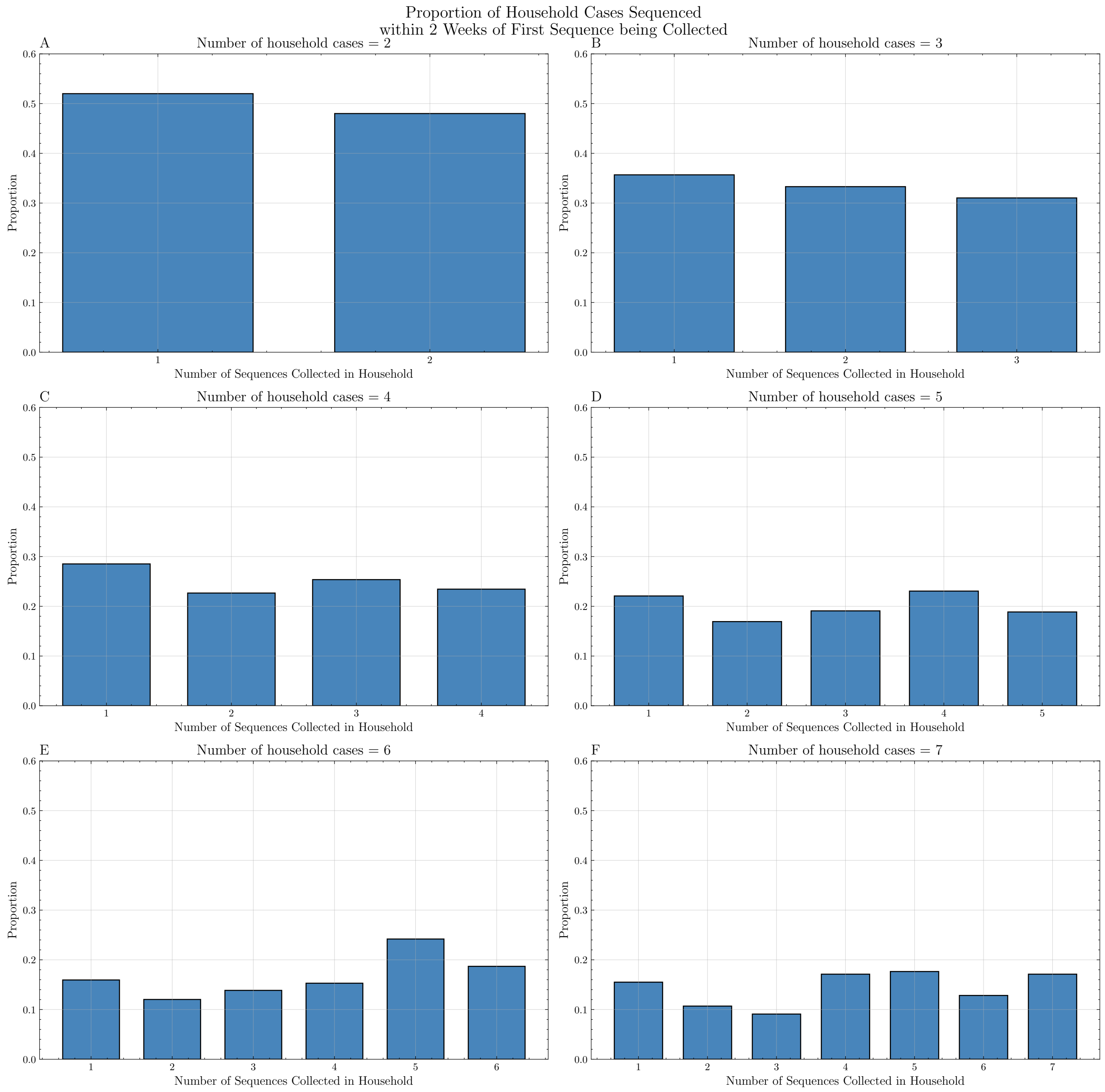


**Supplementary Figure 4.** Proportion of cases in households that return a sequence. Results separated by the number of cases identified within 14 days before or after the first sequence in a household outbreak is collected.

### Appendix 2: Additional Network Results

In this section we present additional analysis and results relating to the construction and structure of the network of plausible transmissions, as well as plausible transmission trees sampled from this network.

Table 1 shows the proportion of edges that are formed by the nation-scale social network that are removed in the process of constructing $G$ due to the genetic difference between the samples being too great, but that would not be removed due to the difference in testing dates. To be more precise, we take the subset of the nation-scale social network $\mathcal{N}$ restricted to the 293,841 samples in our data that gives the household, school, workplace and family relationships between individuals in our data. We then remove all edges for which the time between the first positive tests of connected individuals exceeds 11 days, this being the 95$^{\text{th}}$ percentile of our chosen generation time distribution (see Supplementary Figure 5). This gives a subgraph $\mathcal{N}_g$ of the nation-scale social network, where $g$ denotes the PDF of the generation time distribution. We then calculate the proportion of edges in $\mathcal{N}_g$ for each setting that are not edges in $G$ (since $G$ is also a subgraph of $\mathcal{N}_g$). This gives an idea of how many edges are removed due to the genetic data only, and therefore an idea of how useful the genetic data is in narrowing down the choice of plausible infectors within settings.

Figure 6 shows the emergence and duration of identified settings-based transmission clusters over time, categorised by whether they consist of greater than 5 individuals belonging to schools or workplaces (or both). Each line represents a different cluster. We find, in particular, that the Delta variant is associated with a large number of clusters appearing that are associated with schools.

| **Setting** | **Wildtype (B.1.177)** | **Alpha** | **Eta** | **Delta** | **Omicron (BA.1)** | **All** |
|---|---|---|---|---|---|---|
| Household | 0.959 | 0.950 | 0.986 | 0.797 | 0.933 | 0.429 |
| School | 0.561 | 0.490 | 0.972 | 0.330 | 0.678 | 0.178 |
| Workplace | 0.320 | 0.514 | 0.956 | 0.106 | 0.564 | 0.127 |
| Family | 0.949 | 0.932 | 0.990 | 0.735 | 0.898 | 0.405 |

**Supplementary Table 1.** Proportion of the subset of nation-scale network edges with testing dates less than or equal to 11 days apart (i.e. proportion of edges of $\mathcal{N}_g$) that are also edges in $G$. This is the proportion of possible edges within settings that are not removed by applying our cutoff based on the genetic data.

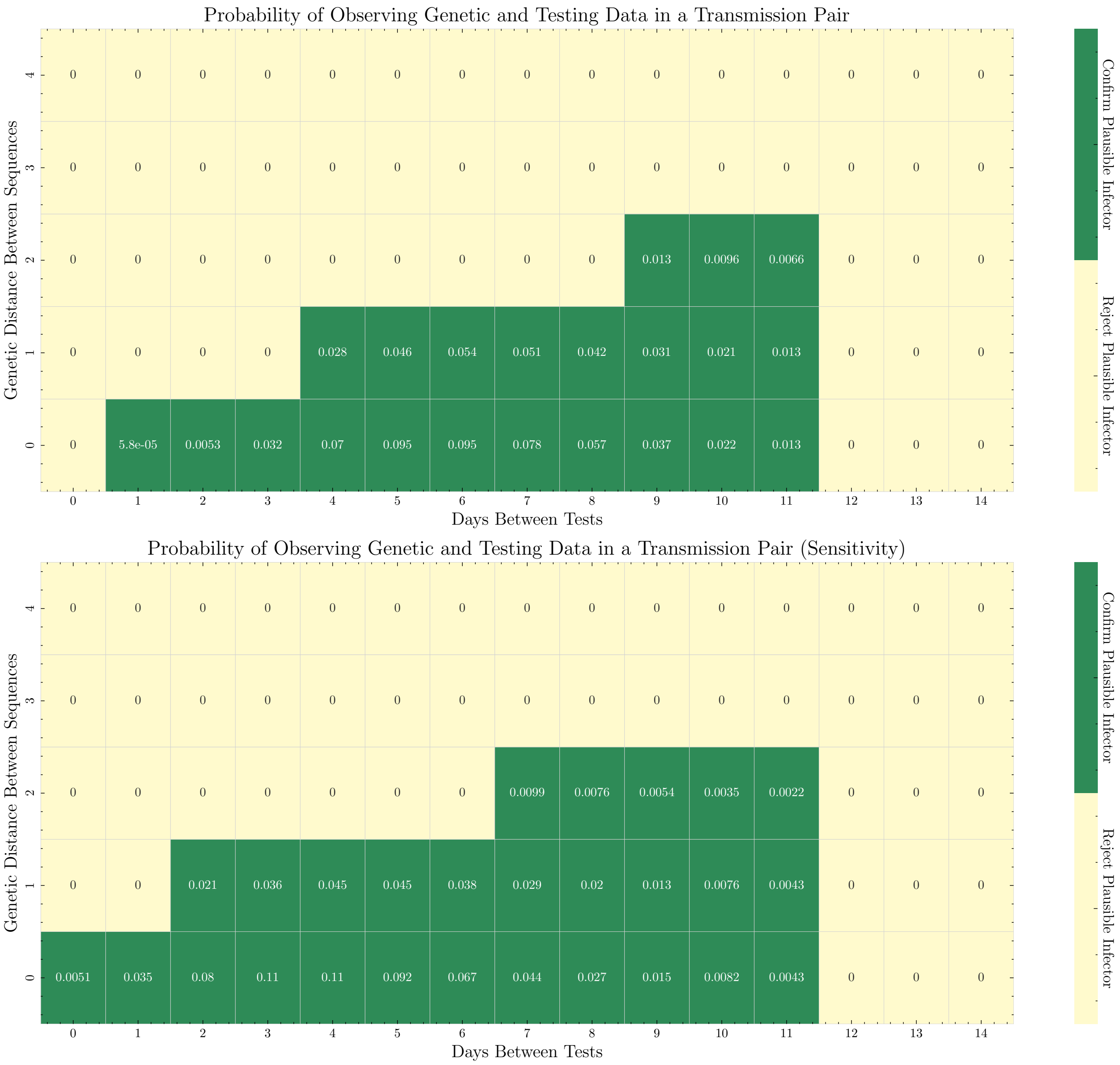


**Supplementary Figure 5.** If two individuals, $A$ and $B$, form a transmission pair, then the joint probability of observing a number of days between the first positive tests from $A$ and $B$, $d$, and of observing a Hamming distance $h$ between the pair of sequences is shown above. The green region defines the region of the probability space that corresponds to accepting the pair $(A, B)$ as a plausible transmission pair. The panel above shows this region for our main analysis, whereas the panel below shows this region for our sensitivity analysis, for which the assumed generation time distribution is shifted to the left by two days.

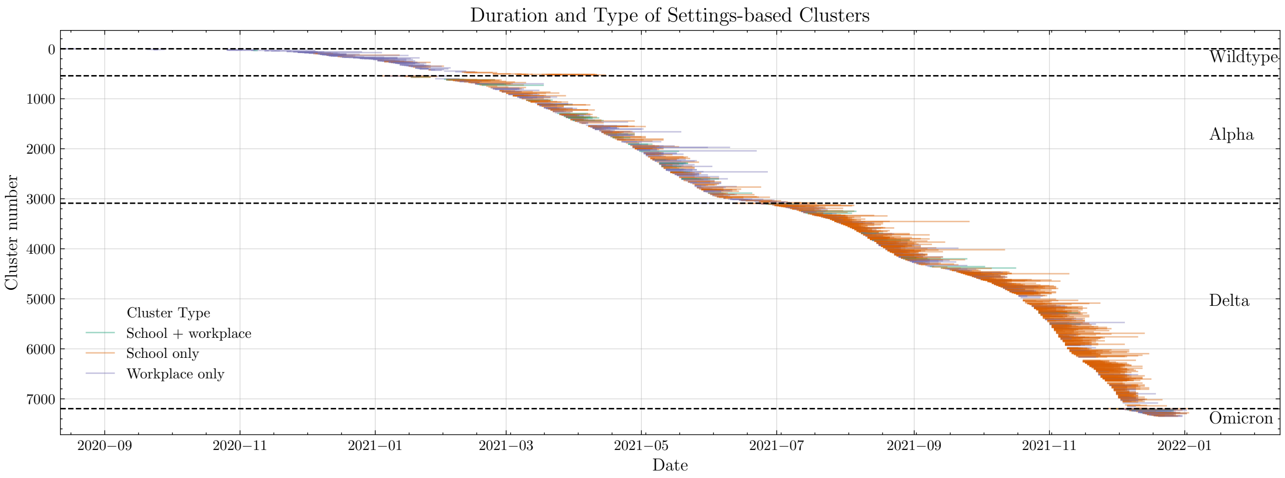


**Supplementary Figure 6.** Durations of clusters for each variant, coloured by type of cluster. If a cluster has more than 5 cases belonging to a school and fewer than 5 belonging to a workplace, then it is denoted as a 'school only' cluster, with 'workplace only' clusters defined analogously. If the cluster contains more than 5 cases of both school and workplace cases, then it is defined as a 'school and workplace' cluster.

### Appendix 3: Sensitivity Analysis of Networks

In this section we present the results of some sensitivity analyses conducted for the construction of the network of plausible transmissions $G$ and show the implications that different choices of strategy for sampling trees can have. In Figure 7 we show the impact of using a shifted generation time distribution on the assignment and weighting of edges in $G$ in terms of the proportion of transmissions that occur in settings over time. This is referred to in the Figure as "Sensitivity Analysis", and results in a slightly higher proportion of transmission pairs identified in settings compared with our standard analysis (panel C of the Figure). In this plot we also demonstrate the impact of the choice of sampling strategy between sampling random trees and prioritised settings trees. Here the difference is pronounced, with random trees seldom choosing transmission pairs within settings when compared with prioritised settings trees. For this reason we choose prioritised settings trees for our main analysis, as we do not believe that the random trees sampling strategy accurately captures the transmission that occurs in settings, giving estimates of transmission that are much lower than other estimates in the literature [12]. Finally, we also include an analysis that gives a small weighting (of $1 \times 10^{-6}$) to pairs with a difference of zero days between their first positive tests, allowing for a serial interval of zero days. This slightly boosts the proportion of transmissions occurring in settings so that it is closer to the proportion obtained in the sensitivity analysis, demonstrating that a number of these pairs are removed in the main analysis due to having an observed serial interval of zero.

Figure 8 shows the proportion of transmissions that occur in educational settings, separated by the type of setting. For our main analysis, we consider only "Grundskole" (Danish compulsory education between the ages of 5 and 16) and "Gymnasium" (Danish secondary education from the age of 16 upwards). This excludes other types of further education including, but not limited to, universities and technical colleges. We show the proportions of plausible transmissions in these kinds of educational settings to transmission in the whole education system in panel B of Figure 8.

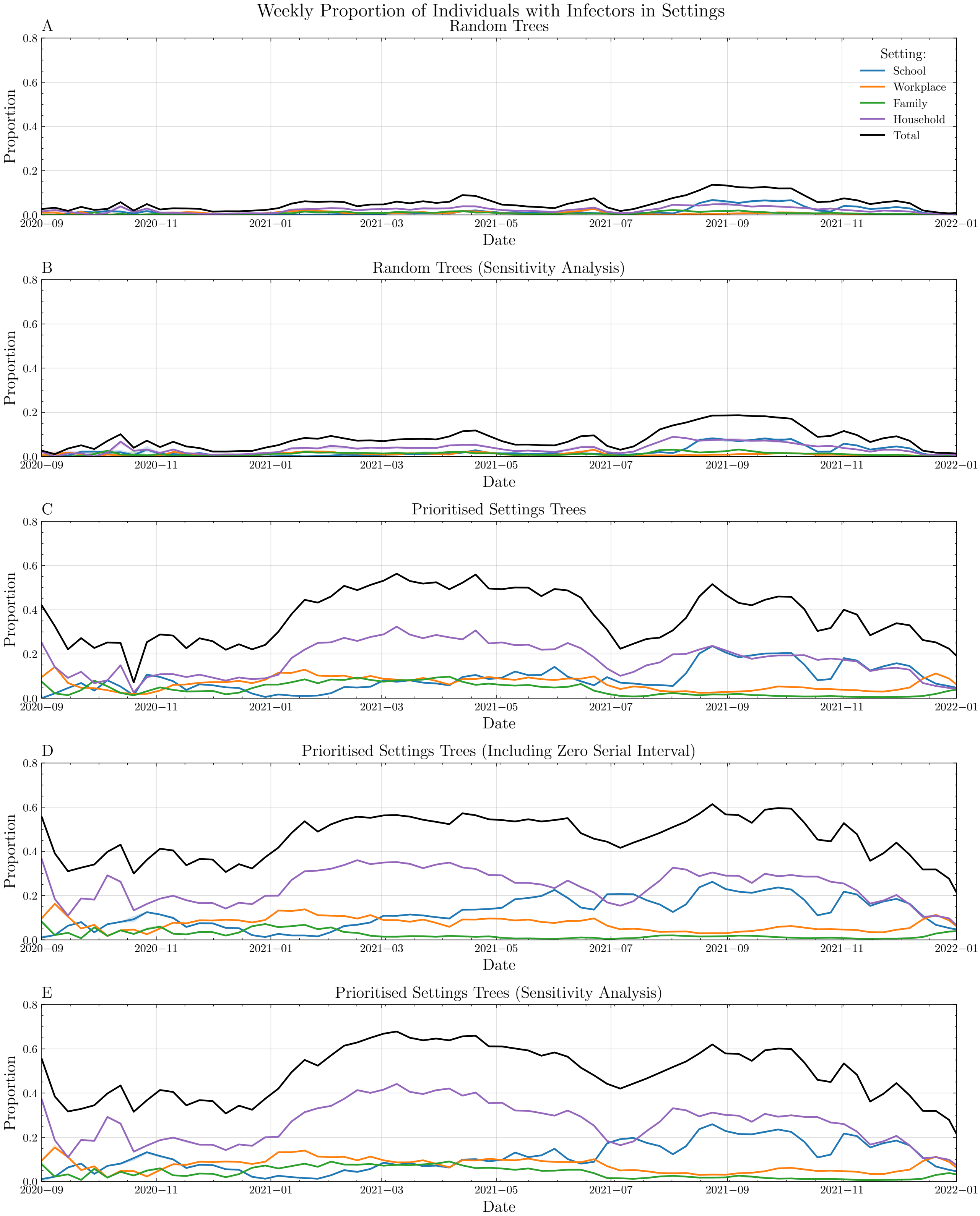


**Supplementary Figure 7.** Comparison of proportions of individuals with a plausible infector in different settings in trees sampled according to different strategies. Panel C corresponds to Figure 2 panel A in the main text).

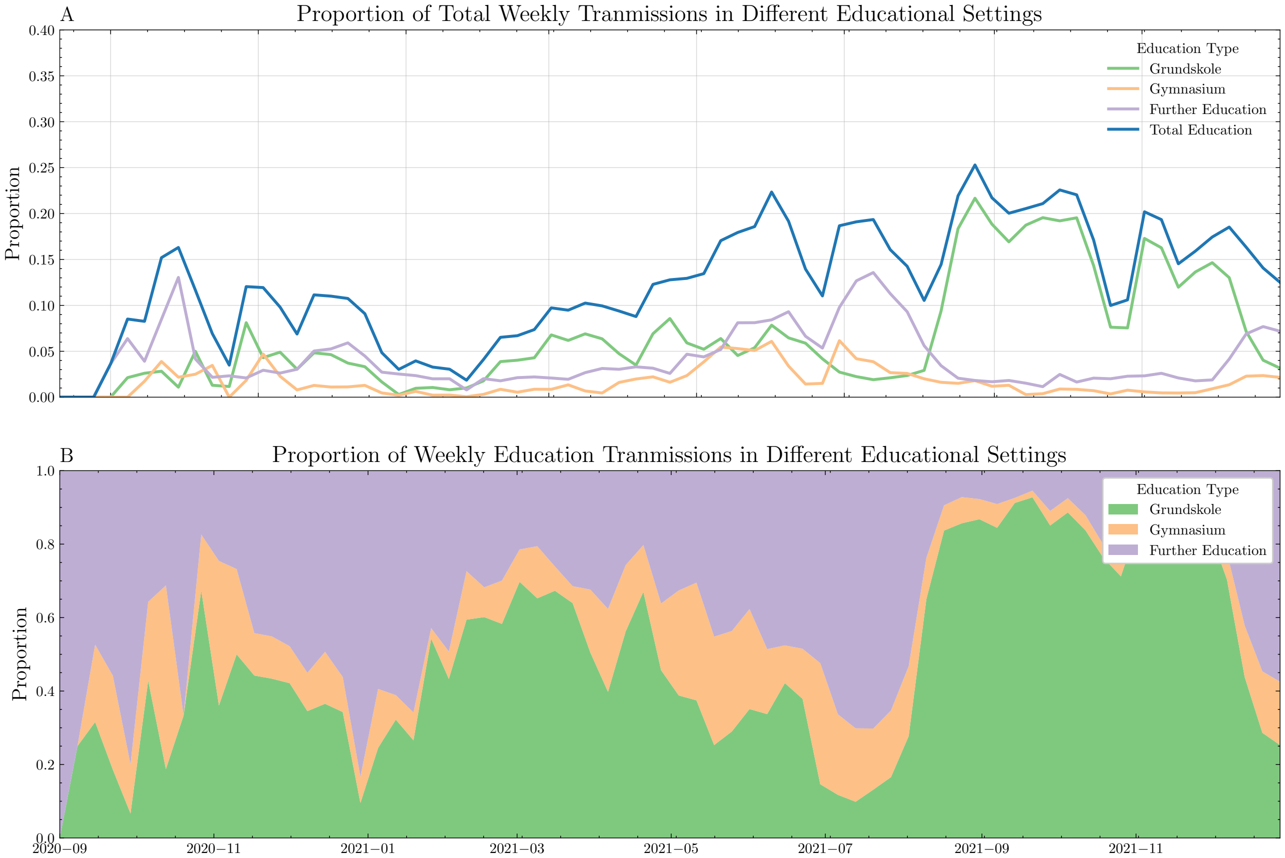


**Supplementary Figure 8.** Proportion of individuals with a plausible infector in an educational institution, separated by type of institution. These proportions are calculated using our baseline model for the generation time interval.

### Appendix 4: NPIs in Denmark

In this section, we provide further details on the NPIs in Denmark, as well as further methods detailing the construction of the instantaneous reproduction numbers used in the main text.

#### 4.1. Further Methods - National aggregated data transmission model

The $R_t$ model we describe here is an extension of the semi-mechanistic branching process model described by Bhatt et al [13]. We are adding a component to the model to infer the time-varying weekly *incidence ascertainment ratios* $(iar)$ following the approach of Mishra et al [14]. We also vary the *infection fatality ratio* $(ifr)$ to account for the dominant variant and vaccination rates. Time is indexed by $t$, ranging from day 1 to $N$, and location, which could be a country, state/region or otherwise defined geographic unit, is indexed by $m$, ranging from location 1 to $M$.

The generation distribution $g$ is unknown but can be approximated by the distribution of the serial interval, the time from onset to onset in an infector-infectee pair. We use a Gamma distribution $g \sim \Gamma(6.5, 0.62)$ which is widely used in the literature, for example, Flaxman et al [15] and the mean is consistent with systematic reviews of the serial interval [16]. We can model the number of infections $i_{t,m}$ for time $t$ and location $m$ using a renewal equation.

$$\begin{aligned} i_{t,m} &= S_{t,m} R_{t,m} \sum_{\tau=0}^{t-1} i_{\tau,m} g_{t-\tau} \\ S_{t,m} &= 1 - \frac{\sum_{j=0}^{t-1} i_{j,m}}{N_m} \end{aligned} \quad (5)$$

We allow for the depletion of susceptible individuals in the population by removing the past infected individuals from the overall population $N_m$ of location $m$. We discretise the generation distribution such that $g_1 = \int_0^{1.5} g(x)dx$ and $g_i = \int_{i-0.5}^{i+0.5} g(x)dx$ for $i \in \mathbb{N} \wedge i > 1$.

$D_{t,m}$ are the number of daily deaths for time $t$ and location $m$ which we model as $d_{t,m} = \mathbb{E}[D_{t,m}]$, that is the expected number of deaths due to COVID-19. We assume that $D_{t,m}$ has a negative binomial distribution with mean and variance as given by equation 6 and $\psi_1$ is a half normal distribution[1].

$$\begin{aligned} D_{t,m} &\sim NegBinomial(d_{t,m}, d_{t_m} + \frac{d_{t,m}^2}{\psi_1}) \\ \psi_1 &\sim N^+(0,5) \end{aligned} \quad (6)$$

Similar to Flaxman et al [15] we link observed deaths and infections using the $ifr_m$, which is different for each location m driven by age structure and other factors for the specific location. The mean estimate $ifr_m$ corresponds to the estimate from early in the pandemic (1.02% for Denmark). We vary this over time as a function of $scale_v$, a multiplicative factor that adjusts the $ifr$ for the dominant SARS-CoV-2 variant. We take this scaling factor relative to the wildtype variant from Perez-Guzman et al. [17] (scaling for $Alpha = 2.42$, $Delta = 1.83$, and $Omicron = 0.58$). We also adjust for the impact of vaccination following the estimates of Watson et al [18] by reducing the effective $ifr$ by 0.35% times the percentage of fully vaccinated people in the population (equation 7). The resultant $ifr$'s for Denmark, by dominant SARS-CoV-2 variant, were $ifr_{WT} = 1.02$, $ifr_{Alpha} = 2.41$, $ifr_{Delta} = 1.62$, and $ifr_{Omicron} = 0.25$. This allows us to capture the benefit of vaccinations indirectly in the model. In addition, we allow for noise around the mean $ifr_m$, since those mean estimates have associated uncertainty. We also require the distribution $\pi$, which is the convolution of the distribution from infection to onset of COVID-19 and the distribution from onset to death of COVID-19, both of which we assume are Gamma distributed, and we take the parameters from the literature [19] and [20], respectively. This allows us to link observed deaths and infections using equation 8 ($\pi$ is discretised in the same way as the generation distribution above).

[1]Let X be normally distributed with zero mean and variance $\sigma$ and define $Y = |X|$ then X has a half positive normal distribution with zero mean and variance $\sigma$, that is $Y \sim N^+(0, \sigma)$.

$$\begin{aligned} ifr^*_m &\sim ifr_{m,v,t} \cdot N(1, 0.1) \\ ifr_{m,v,t} &= ifr_m * scale_v - 0.0035 * vacc_t \quad \textbf{(7)} \\ \pi &\sim \Gamma(5.1, 0.86) + \Gamma(17.8, 0.45) \\ d_{t,m} &= ifr^*_m \sum_{\tau=0}^{t-1} i_{\tau,m}\pi_{t-\tau} \quad \textbf{(8)} \end{aligned}$$

We also observe daily reported COVID-19 cases $C_{t,m}$ which are modelled as $c_{t,m} = \mathbb{E}[C_{t,m}]$. Analogously to deaths we assume that $C_{t,m}$ has a negative binomial distribution as given by equation 9. To link infections and cases we follow the approach of Mishra et al [14] who assume that the distribution $\pi^{i2c}$, the time lag between infection and case identification, is zero for the first 3 days and 10% for the subsequent 10 days. This allows us to link observed cases with infections and infer the incident ascertainment rate ($iar$). We specify the $iar$, as given in Equation 11, as a weekly random walk (we provide details on this below) with link function $f(x) = 2 \cdot \frac{e^x}{1+e^x}$, which is a twice inverse logit function.

$$\begin{aligned} C_{t,m} &\sim NegBinomial(c_{t,m}, c_{t_m} + \frac{c^2_{t,m}}{\psi_2}) \quad \textbf{(9)} \\ \psi_2 &\sim N^+(0, 5) \\ \pi^{i2c} &= c(0, 0, 0, repeat(\frac{1}{10}, 10)) \\ c_{t,m} &= \sum_{\tau=0}^{13} iar_{w(t),m} i_{\tau,m} \pi^{i2c}_{t-\tau} \quad \textbf{(10)} \\ iar_{w(t),m} &= f(-\varepsilon_{w_m(t),m}) \quad \textbf{(11)} \end{aligned}$$

The time varying reproduction number $R_{t,m}$ for time $t$ and location $m$ can be expressed as linear function of covariates with the same link function as previously specified. $X_t, m, k$ are state specific covariates which have the same estimate across all locations (fixed effect). The priors for the fixed effect coefficients are $\alpha_k \sim N(0, 0.5)$. We could also allow for additional effects for groups of locations (regions of locations) in this model. $Z_t$ is the covariate to allow us to estimate location specific (random) effects. $\epsilon_{w_m(t),m}$ is a random walk which captures residual variation not explained by the covariates. We specify the random walk for $\epsilon_{w_m(t),m}$ such that it is equivalent to a second order autoregressive process AR(2), which has mean zero. We parameterise this AR(2) process to be weekly. We take a prior for the basic reproduction number as $R_{0,m} \sim N(3.28, \kappa)$, where $\kappa \sim N^+(0, 0.5)$ [21].

$$R_{t,m} = R_{0,m} \cdot f\Big(-\sum X_{t,m,k}\alpha_k - Z_{t,m}\alpha_m^{location} - \epsilon_{w_m(t),m}\Big) \quad \textbf{(12)}$$

One of the challenges is to find the actual starting point of the pandemic and when to start seeding new infections. Following Mishra et al [14] we start to seed infections 30 days prior to the cumulative tenth observed death in that location. Following that date we see the model on 6 consecutive days where $i_{j,m} \sim Exponential(1/\tau)$ for $j = 1, \cdots, N0$ with $N0 = 6$ and $\tau \sim Exponential(0.03)$.

In the above model we have two random walks $\varepsilon_{w_m(t),m}$ and $\epsilon_{w_m(t),m}$ for the $iar$ and $R_t$ respectively. Both are weekly AR(2) processes but with different parameterisations. We use a weekly process to reduce noise and provide more stability in our inferred parameters. We assume that $\varepsilon_{w_m(t),m}$ has mean 0.4 (corresponding to a 40% $iar$ at the beginning of the pandemic) and variance $(\sigma^*_{w,iar})^2$, which is different for each location $m$, and starts at the beginning of the seeding period. The autoregressive coefficients of the AR(2) process have priors $\rho_{1,iar}$ and $\rho_{1,iar}$ as defined in equation 13 and we impose the condition that the $\rho_{i,iar} \in [0, 1]$. Similarly we define the random walk $\varepsilon_{w_m(t),m}$ for $R_t$ as an AR(2) with zero mean and variance $(\sigma^*_w)^2$. The AR(2) coefficients have the same constraints $\rho_i \in [0, 1]$.

$$
\begin{aligned}
\varepsilon_{1,m} &\sim N(0.4, 0.01) & \epsilon_{1,m} &\sim N(0, 0.01) \\
\varepsilon_{w,m} &\sim N(\rho_{1,iar}\varepsilon_{w-1,m} + \rho_{2,iar}\varepsilon_{w-2,m}, (\sigma^*_{w,iar})^2) & \epsilon_{w,m} &\sim N(\rho_1\epsilon_{w-1,m} + \rho_2\epsilon_{w-2,m}, (\sigma^*_w)^2) \\
\rho_{1,iar} &\sim N(0.4, 0.1) & \rho_1 &\sim N(0.8, 0.05) \\
\rho_{2,iar} &\sim N(0.2, 0.05) & \rho_2 &\sim N(0.1, 0.05) \\
\sigma_{w,iar} &\sim N^+(0, 0.1) & \sigma_w &\sim N^+(0, 0.2)
\end{aligned} \tag{13}
$$

We need one additional constraint on the standard deviation for both random walks to ensure stationarity of the AR(2) process, which is given by equation 14. Equation 15 provides the translation form daily to weekly frequency, $w_{m(t)}$ is kept constant for one week before being incremented, starting at the beginning of the seeding period.

$$
\sigma^*_w = \sigma_w\sqrt{1 - \rho_1^2 - \rho_2^2 - 2\rho_1^2\rho_2/(1-\rho_2)} \tag{14}
$$

$$
w_m(t) = \left\lfloor \frac{t - t_m^{start}}{7} \right\rfloor + 1 \tag{15}
$$

We have set out the model here, including covariates in equation 12. We can also run this model without any covariates and just a random walk, as we will see in the results section.

The parameters of this model are jointly estimated, and the inference was performed in R using Stan [22]. In the implementation, we made a small adjustment to equation 8 for computational efficiency. Usually, this sum goes back to $t-1$, that is, the full history. For short runs, this is feasible; however, for longer runs, such as 2 years in our case, this becomes computationally problematic. We restrict equation 8 to a maximum look back of 71, which covers 99.98% of the density of $\pi$ and does not impact the results.

***4.1.1. Instantaneous reproduction number results and comparison.*** In Figure 9, we provide the results of the $R_t$ model for Denmark for 2020/21. This model did not include any covariates $X_{t,m,k}$ and was calibrated to reported deaths. A timeline of the COVID-19 pandemic in Denmark, compiled by the Statens Serum Institut, outlining the key events and restrictions, is available at [23].

Panel (A) displays the number of daily reported cases (brown bars) and the inferred number of infections (blue ribbon). The first case was reported on 27 February 2020, the first restrictions were introduced on 3 March 2020, and cases increased rapidly in early March. The peak of inferred infections during the first wave was on 11 March with a median estimate of 15,388 infections. Reported cases remained low for most of 2020 and began to increase significantly in October, peaking on December 18, 2020, at 4,508. Inferred infections peaked on 23 December 2020 at a median estimate of 42,075 before restrictions became effective, and both reported cases and inferred infections declined. The number of reported cases, as well as the inferred number of infections, remained low until October 2021, rarely exceeding 1,000. From November 2021, reported cases increased steadily, and this trend accelerated with the arrival of the Omicron variant in Denmark in December, resulting in daily reported cases exceeding 20,000.

Panel (B) displays the number of daily reported deaths used to fit the model. The first reported death occurred on 13 March 2020, and the peak of the first wave was on 4 April 2020 with 22 deaths. In line with reported cases, deaths remained low throughout the spring, summer, and autumn. Reported deaths increased over November and December 2021, but remained at levels below the peak in December 2020/January 2021 of 37 deaths (the report of 60 deaths on 15 January 2021 was across 2 days), as a large part of the Danish population had received a full course of vaccination. Most of the vaccination campaign took place in the first half of 2021, leading to waning immunity against infection and accounting for the large number of reported cases and inferred infections. However, protection against death remained high, which explained the low number of reported deaths.

In panel (C), we display the estimated instantaneous weekly reproduction numbers $R_t$. The initial estimate of $R_t$ was 3.10, but with a wide credible interval (95% CrI: 2.12 to 4.18). $R_t$ declined rapidly with the introduction of NPIs and was below one by the end of March 2020 (0.78, 95% CrI: 0.55 to 1.05)[2]. $R_t$ increased above one again over the summer

[2] The 95$^{th}$ percentile of the confidence interval was below one by 2 April 2020.

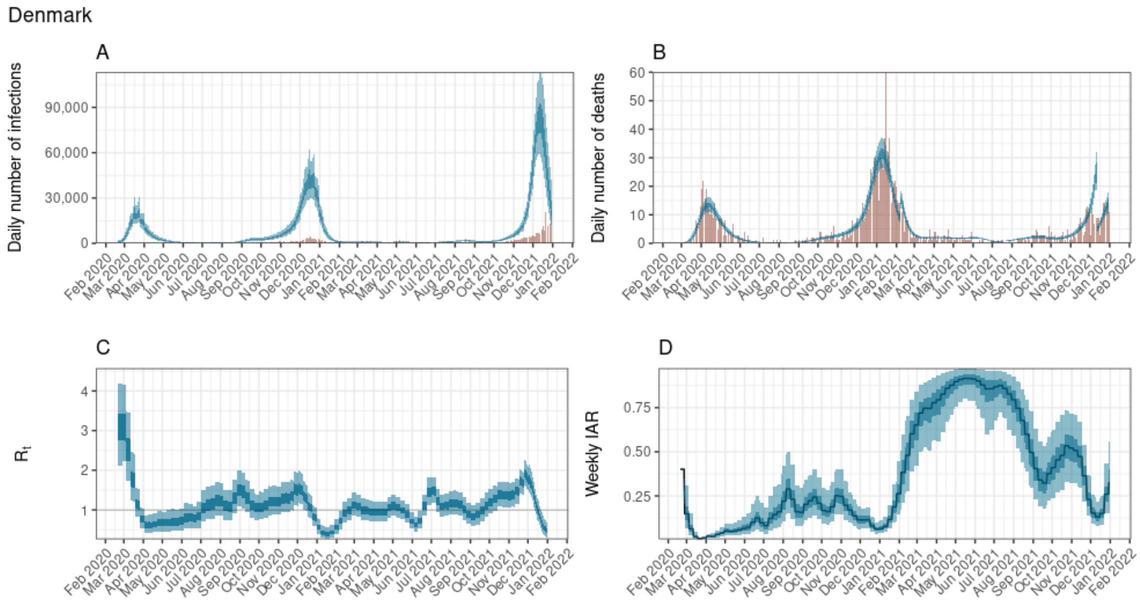


**Supplementary Figure 9.** $R_t$ model for Denmark: The black line indicates the central estimate, the dark blue ribbon indicates the 50% credible interval, and the light blue ribbon indicates the 95% credible interval for all plots. (A) daily number of infections with reported cases displayed as coral coloured bars (B) daily number of deaths with reported deaths displayed as coral coloured bars (C) $R_t$ estimate (D) incident ascertainment ratio.

and at the start of the new school year, but at a low number of reported cases and inferred infections. By the beginning of December 2020, $R_t$ peaked at 1.55 (95% CrI: 1.16 to 1.99), stayed at this elevated level for most of December before dropping below one at the beginning of January 2021. During the first quarter of 2021, $R_t$ remained very low as a range of NPIs were in place and only started to increase as those measures were lifted. Over the summer of 2021, $R_t$ remained close to 1 and increased again at the end of the summer holidays as schools returned (without any restrictions). At the end of 2021, $R_t$ increased again with the arrival of the Omicron variant in Denmark.

Panel (D) displays the inferred weekly infection ascertainment ratios ($iar$). Similar to most of Europe, the $iar$ was very low during the first 6 months of the pandemic, as testing capacity was limited. Over the summer of 2020, the $iar$ fluctuated around 0.25 due to a low number of reported cases and inferred infections. During the Christmas wave of 2020, $iar$s were low as testing had not been expanded to meet demand. The Danish government increased PCR testing and sequencing hugely in January 2021. This enabled Denmark to capture a large proportion of infections as reported cases, and the $iar$ remained above 0.5 for large parts of 2021.

We compare $R_t$ estimates from the aggregated data model to $R_t$ estimates from the LSHTM model `EpiNow2` [24]. Overall, the estimates are consistent and the 90% credible intervals overlap. One exception is January 2021, when the `EpiNow2` estimates are higher but still significantly below 1, before returning to line by the end of February. This difference is most likely due to the fact that `EpiNow2` is fit only to reported cases.

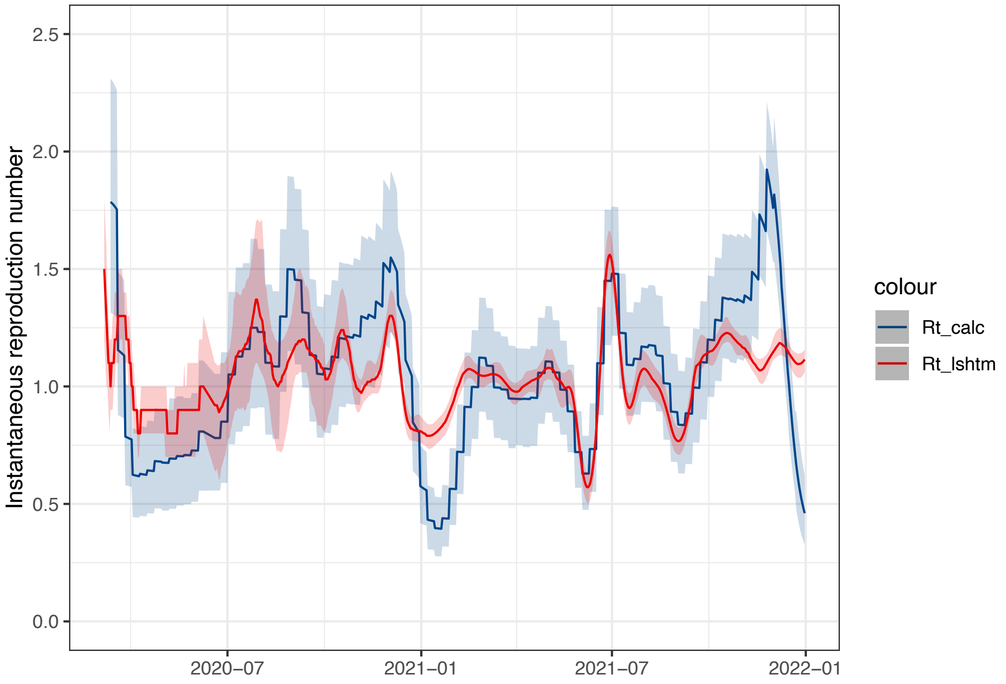


**Supplementary Figure 10.** $R_t$ model output comparison between the aggregated data model(blue) and LSHTM EpiNow2 model [24] (red). The coloured ribbons indicated 90% credible intervals.

##### 4.2. School Restrictions

A range of restrictions in schools was in place in Denmark over the first two years of the pandemic. In this section, we provide an overview based on the summary provided by Danmarks Evalueringsinstitut [25], the Danish Evaluation Institute, which is part of the Danish Ministry of Children and Education.

It is important to note that although the rules talk about primary school in Denmark, this includes secondary education up to the age of 16. In the UK context, for example, this would be education up to and including GCSEs. Education up to grade 9 is compulsory, and grade 10 is optional.

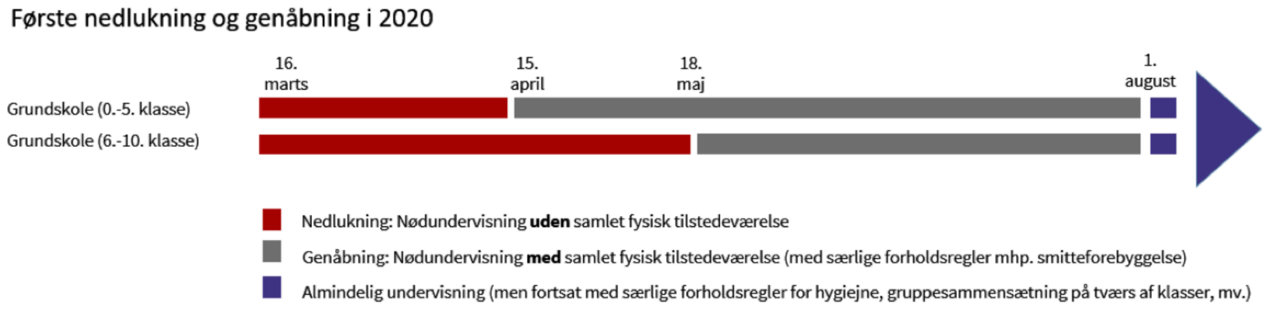


**Supplementary Figure 11.** School restrictions in place over 2020 in Denmark [25] (red = no physical attendance, grey = emergency teaching with precautions, dark blue = normal teaching with precautions).

###### *4.2.1. First lockdown and reopening in 2020.* Primary school (grades 0–5)

- *16 Mar*: Closure: Emergency teaching without any physical attendance (red)
- *15 Apr*: Reopening: Emergency teaching with physical attendance (with special precautions for infection prevention) (grey)
- *18 May*: Continued emergency teaching with precautions (grey)

- *1 Aug*: Return to normal teaching (still with hygiene rules, grouping precautions) (dark blue)

**Primary school (grades 6–10)**

- *16 Mar*: Closure: Emergency teaching without physical attendance (red)
- *18 May*: Reopening: Emergency teaching with physical attendance (grey)
- *1 Aug*: Normal teaching (dark blue)

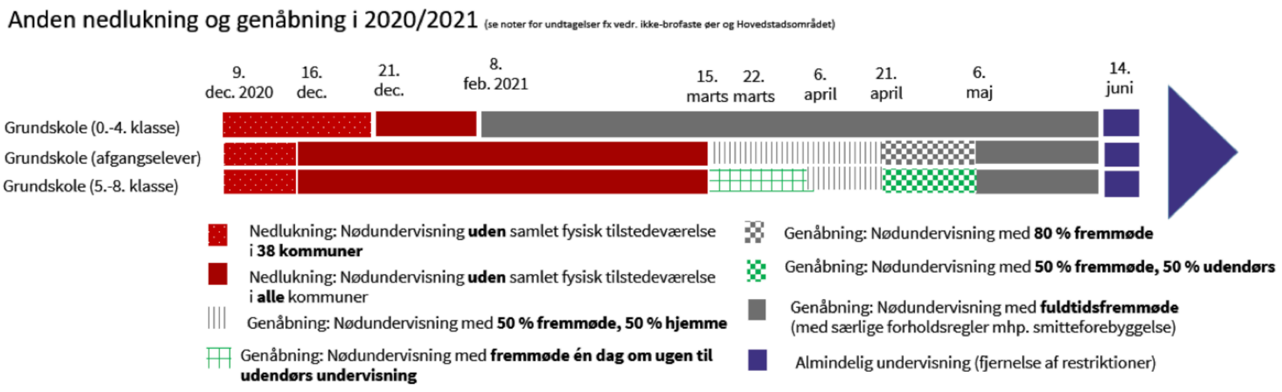


**Supplementary Figure 12.** School restrictions in place over 2021 in Denmark [25].

***4.2.2. Second lockdown and reopening in 2020/2021.*** See notes 1-3 below for exceptions to these rules, e.g., non-bridged islands and the Capital Region.

**Primary school (grades 0–4)**

- *9 Dec 2020*: Closure: Emergency teaching without attendance in 38 municipalities (red with dots)
- *21 Dec 2020*: Closure in all municipalities (solid red)
- *8 Feb 2021*: Reopening: Emergency teaching with 100% attendance (grey)
- *15 Mar 2021*: Normal teaching (grey)
- *14 Jun 2021*: Removal of restrictions → Normal teaching (dark blue)

**Primary school (graduating classes)**

- *9 Dec 2020*: Closure: Emergency teaching without attendance in 38 municipalities (red with dots)
- *16 Dec 2020*: Closure in all municipalities (solid red)
- *8 Feb 2021*: Closure continues (solid red)
- *16 Mar 2021*: Partial reopening: Emergency teaching with 50% attendance, 50% at home (white with grey stripes)
- *21 Apr 2021*: Reopening: Emergency teaching with 80% attendance (grey crosshatch)
- *6 May 2021*: Reopening: Emergency teaching with full attendance (grey)
- *14 Jun 2021*: Removal of restrictions → Normal teaching (dark blue)

**Primary school (grades 5–8)**

- *9 Dec 2020*: Closure: Emergency teaching without attendance in 38 municipalities (red with dots)

- *16 Dec 2020*: Closure in all municipalities (solid red)
- *8 Feb 2021*: Closure continues (solid red)
- *15 Mar 2021*: Reopening: Emergency teaching with outdoor teaching one day per week (green crossed lines)
- *6 Apr 2021*: Reopening: Emergency teaching with 50% attendance, 50% at home (white with grey stripes)
- *21 Apr 2021*: Reopening: Emergency teaching with 50% attendance, 50% outdoors (white with green crosshatch)
- *6 May 2021*: Reopening: Emergency teaching with full attendance (grey)
- *14 Jun 2021*: Removal of restrictions → Normal teaching (dark blue)

**Notes 1:** Figure 12 reflects the main time points of the national closure and reopening periods in 2020 and 2021. Students in special-needs classes and schools have been exempted from being sent home during the second national closure of schools. Vulnerable students were offered emergency education in schools, particularly during the second closure and reopening period. There have also been several local closures and reopenings as part of the circuit breaker municipality closures [26]. As a result, the extent of emergency education without physical attendance has been more extensive than the figure depicts in some areas and schools, due to local and regional closures. These exceptions are further described below.

**Notes 2:** *Closure in December 2020:* Students in grades 5-10 were sent home in 38 municipalities as early as 9 December 2020. As of 16 December 2020, this was expanded to all municipalities. As of 21 December 2020, the program was expanded to all students in grades 0-4 in all municipalities. Students in grades 5-8 at schools in North Jutland were sent home from November 9 to 16, 2020.

**Notes 3:** *Reopening in February-May 2021:* Leaving pupils (graduating classes in 9th and 10th grade), depending on the region, first returned with 50% attendance on 15 and 22 March 2021, respectively. On islands not connected by bridge, pupils in 5th-8th grade and leaving pupils returned with 100% attendance on 1 March 2021. Fifty per cent attendance for 5th-8th grade was introduced nationally (except Brøndby, Ishøj, Vallensbæk, and Høje-Taastrup municipalities) on April 6. On 15 March, pupils in 5th-8th grade in all regions were allowed to attend outdoor classes one day a week. On 21 April, pupils graduating from primary schools were allowed to attend with 80% attendance. However, this did not apply in the Capital Region, specifically in Copenhagen city, the Copenhagen area, North Zealand, and East Zealand. Here, 50% attendance for graduating students continued. Students in grades 5-8 were allowed to attend outdoors during the weeks when they could not attend classes in person – i.e., they had 50% indoor classes and 50% outdoor classes.

#### 4.3. Oxford COVID-19 Government Response Tracker

Data for restrictions were obtained from the Oxford COVID-19 Government Response Tracker [27, 28] and are displayed in Figure 13. The behavioural survey data was obtained from [29].

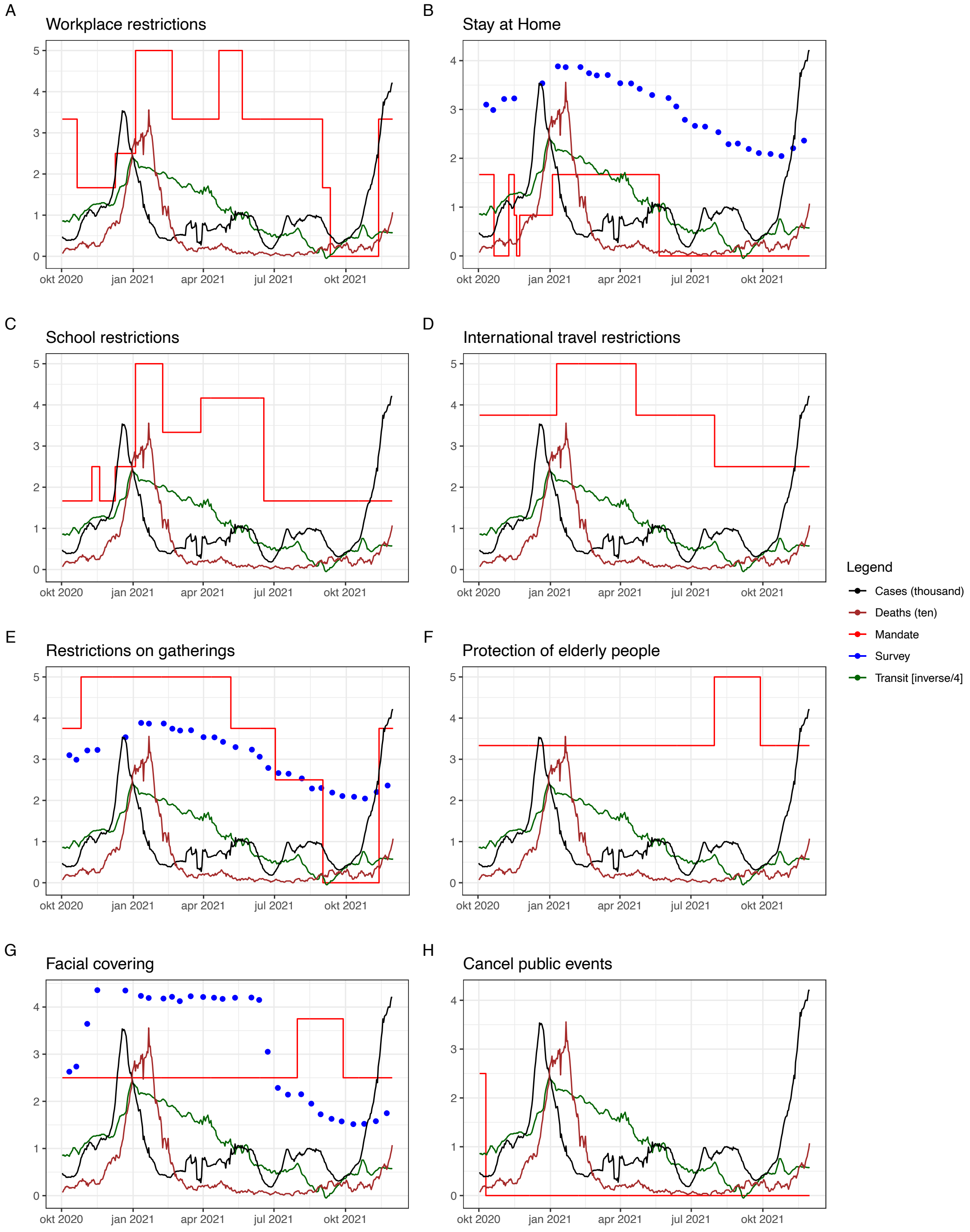


**Supplementary Figure 13.** Time series of NPIs in place from Oct 2020 to Dec 2021 in Denmark. Red lines indicate NPI restriction levels (rescaled to a scale between 0 and 5 for display purposes only, in the model, NPIs are rescaled 0 to 1), black lines display reported, national cases (in thousands), brown lines report deaths (in tens), green lines display the transit index for mobility (inverted and scaled by a factor of 0.25), and the blue dots display survey results ranging from 1 (low adherence) to 5 (high adherence)

###### *4.3.1. Restriction Codebook.*

| ID | Name | Description | Coding |
|---|---|---|---|
| C1 | School closing | Record closings of schools and universities | 1 - recommend closing or open with alterations<br>2 - require closing (only some levels or categories)<br>3 - require closing all levels |
| C2 | Workplace Closure | Record closings of workplaces | 1 - recommend closing or all businesses open with changes<br>2 - require closing (only some levels or categories)<br>3 - require closing for all-but-essential workplaces |
| C3 | Cancel public events | Record cancelling public events | 1 - recommend cancelling<br>2 - require cancelling |
| C4 | Restrictions on gatherings | Record limits on gatherings | 1 - restrictions above 1000 people<br>2 - restrictions between 101-1000 people<br>3 - restrictions between 11-100 people<br>4 - restrictions on 10 people or less |
| C5 | Close public transport | Record closing of public transport | 1 - recommend closing<br>2 - require closing |
| C6 | Stay at home requirement | Record orders to "shelter-in-place" and otherwise confine to the home | 1 - recommend not leaving house<br>2 - require not leaving house with exceptions |
| C7 | Restrictions on internal movement | Record restrictions between cities/regions on internal movement | 1 - recommend not to travel between regions/cities<br>2 - internal movement restrictions in place<br>3 - require not leaving house with minimal exceptions |
| C8 | International travel controls | Record restrictions on international travel [a] | 1 - screening arrivals<br>2 - quarantine arrivals from some or all regions<br>3 - ban arrivals from some regions<br>4 - ban on all regions or total border closure |

**Supplementary Table 2.** OxCGRT Codebook Containment and closure policies [27, 28]: Missing data will be represented as blank in the database, Coding 0 will be applied if no measure was in place at that moment in time. All Measures are at Ordinal Scale.

[a]Note: this records policy for foreign travellers, not citizens

| ID | Name | Description | Measure | Coding |
|---|---|---|---|---|
| E1 | Income Support | Record if the government is providing direct cash payments to people who lose their jobs or cannot work.[a] | Ordinal scale | 1 - government is replacing less than 50% of lost salary (or if a flat sum, it is less than 50% median salary)<br>2 - government is replacing 50% or more of lost salary (or if a flat sum, it is greater than 50% median salary) |
| E2 | Debt/contract relief | Record if the government is freezing financial obligations for households | Ordinal scale | 1 - narrow relief, specific to one kind of contract<br>2 - broad debt/contract relief |
| E3 | Fiscal measures | Announced economic stimulus spending | USD | Record monetary value in USD of fiscal stimuli, includes any spending or tax cuts NOT included in E4, H4 or H5 |
| E4 | International support | Announced offers of COVID-19 related aid spending to other countries | USD | Record monetary value in USD |

**Supplementary Table 3.** OxCGRT Codebook economic policies [27, 28]: Missing data will be represented as blank in the database, Coding 0 will be applied if no measure or support was in place at that moment in time.

[a]Note: only includes payments to firms if explicitly linked to payroll/salaries

| ID | Name | Description | Measure | Coding |
|---|---|---|---|---|
| H1 | Public information campaigns | Record presence of public info campaigns | Ordinal scale | 1 - public officials urging caution about Covid-19<br>2 - coordinated public information campaign |
| H2 | Testing policy | Record government policy on who has access to testing | Ordinal scale | 1 - only those who both (a) have symptoms AND (b) meet specific criteria<br>2 - testing of anyone showing Covid-19 symptoms<br>3 - open public testing |
| H3 | Contact tracing | Record government policy on contact tracing after a positive diagnosis [a] | Ordinal scale | 1 - limited contact tracing; not done for all cases<br>2 - comprehensive contact tracing; done for all identified cases |
| H4 | Emergency investment in healthcare | Announced spending on healthcare system[b] | USD | Record monetary value in USD |
| H5 | Investment in vaccines | Announced public spending on Covid-19 vaccine development | USD | Record monetary value in USD |
| H6 | Facial coverings | Record policies on the use of facial coverings outside the home | Ordinal scale | 1 - Recommended<br>2 - Required in some specified shared/public spaces<br>3 - Required in all shared/public spaces<br>4 - Required outside the home at all times |
| H7 | Vaccination policy | Record policies for vaccine delivery for different groups | Ordinal scale | 1 - Availability for ONE group[c]<br>2 - Availability for TWO groups<br>3 - Availability for ALL groups<br>4 - Available to ALL groups + some others<br>5 - Universally available |
| H8 | Protection of elderly people | Record policies for protecting elderly people in LT Care Facilities/community setting | Ordinal scale | 1 - Recommended restriction measures in LTCFs<br>2 - Narrow restrictions measures in LTCFs<br>3 - Extensive restrictions measures in LTCFs |

**Supplementary Table 4.** OxCGRT Codebook Health system policies [27, 28]: Missing data will be represented as blank in the database, Coding 0 will be applied if no measure was in place at that moment in time.

[a]Note: we are looking for policies that would identify all people potentially exposed to Covid-19; voluntary bluetooth apps are unlikely to achieve this
[b]Note: only record amount additional to previously announced spending
[c]Groups: key workers/ clinically vulnerable groups (non elderly) / elderly groups

### Appendix 5: Reproduction numbers and NPI further results

In this section, we provide an extended set of results for the model assessing the impact of NPIs, based on individual-level data, presented in the main text.

#### 5.1. Case reproduction number model fits

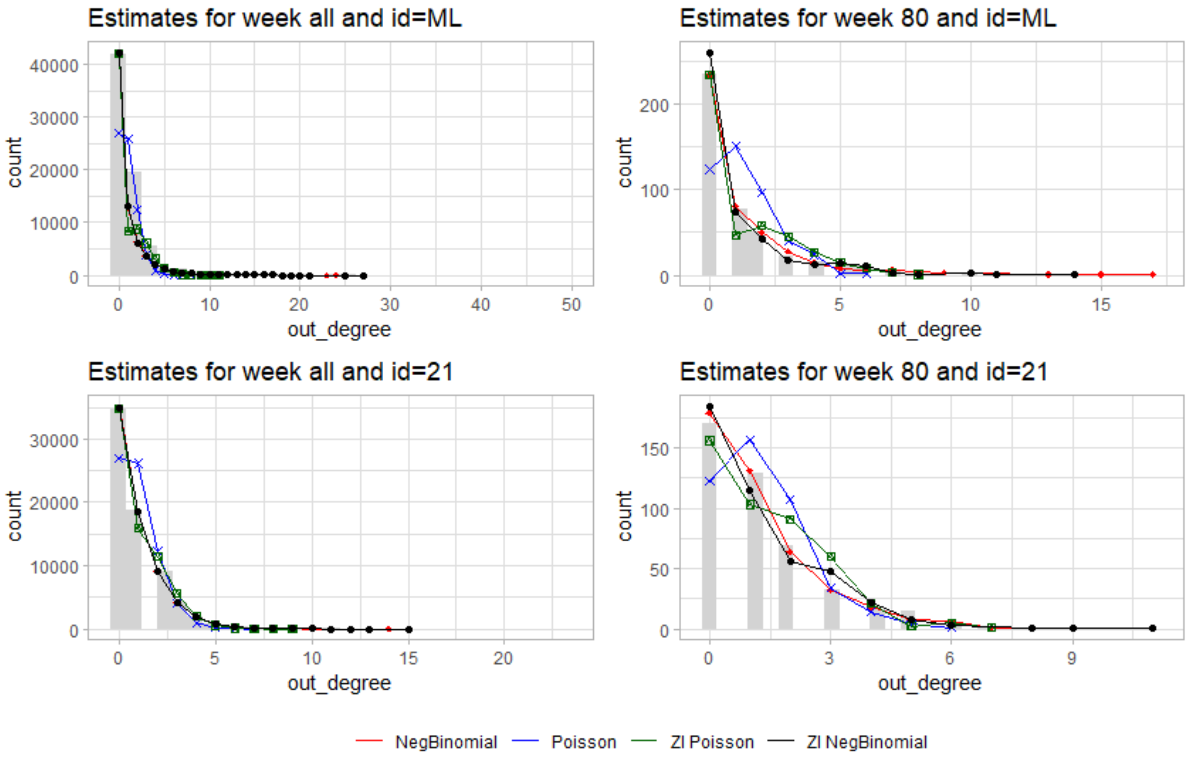


**Supplementary Figure 14.** Effective reproduction numbers and overdispersion: (A) Comparison of aggregate, adjusted $R_t$ with case reproduction numbers based on random trees and prioritised trees, (B) Comparison of overdispersion estimates based on random and prioritised trees, (C) Decomposition of prioritised settings case reproduction number by setting, (D) Overdispersion estimate for prioritised setting for households (beta binomial model) and schools, workplaces and the community (negative binomial model), (E) Decomposition of random trees case reproduction number by setting, and (F) Overdispersion estimate for random trees for households (beta binomial model) and schools, workplaces and the community (negative binomial model).

#### 5.2. Extended main text results, including Random Trees

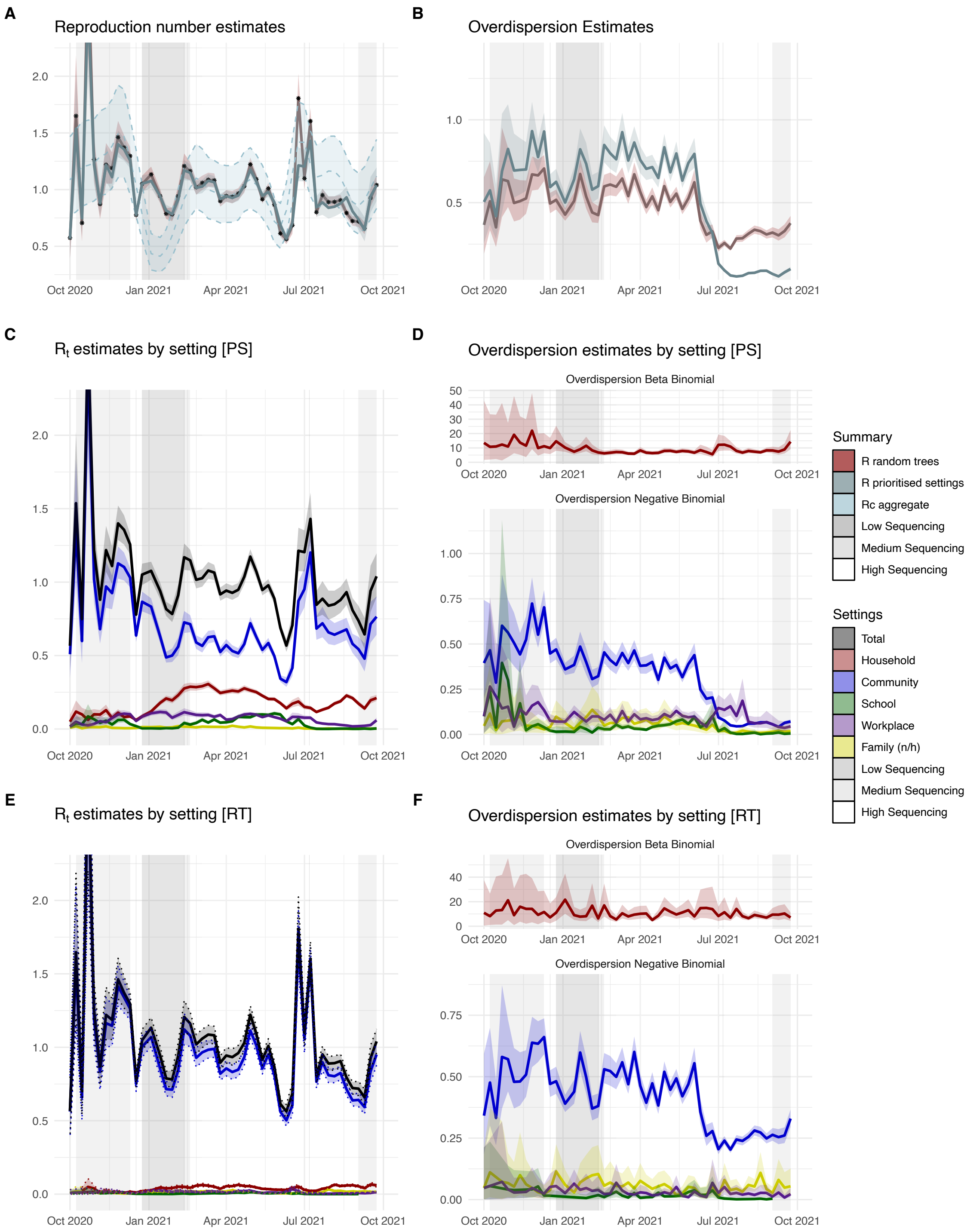


**Supplementary Figure 15.** Effective reproduction numbers and overdispersion: (A) Comparison of aggregate, adjusted $R_t$ with case reproduction numbers based on random trees and prioritised trees, (B) Comparison of overdispersion estimates based on random and prioritised trees, (C) Decomposition of prioritised settings case reproduction number by setting, (D) Overdispersion estimate for prioritised setting for households (beta binomial model) and schools, workplaces and the community (negative binomial model), (E) Decomposition of random trees case reproduction number by setting, and (F) Overdispersion estimate for random trees for households (beta binomial model) and schools, workplaces and the community (negative binomial model).

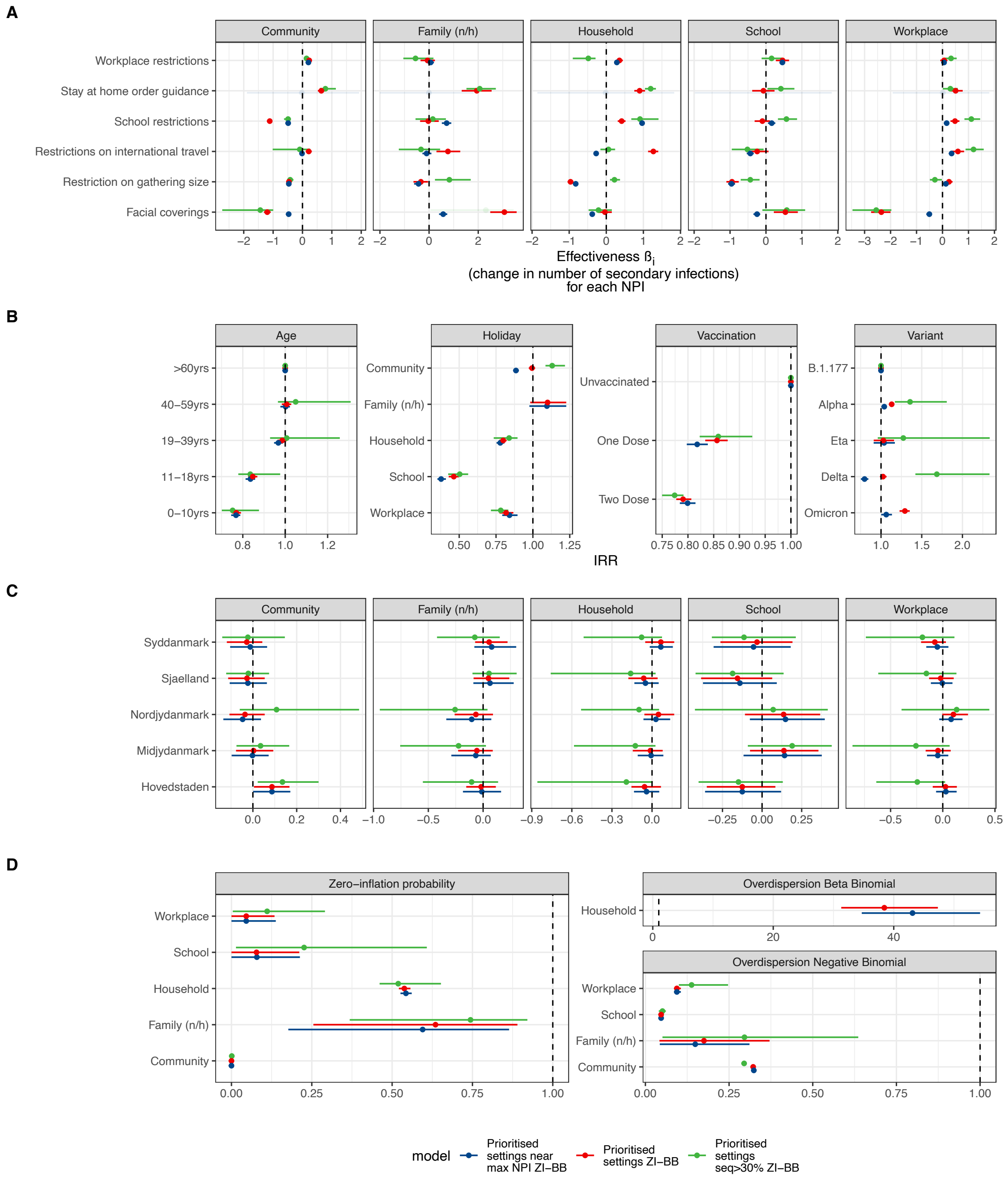


**Supplementary Figure 16.** Effectiveness of NPIs based on individual level data: (A) Effect sizes of association between number of infections caused by an infector and level of each NPI by setting. (B) Risk heterogeneities of the number of secondary infections by Age, School holidays, Vaccination status and SARS-CoV-2 variant. (C) Regional random effects estimates by setting. (D) Zero-inflation and overdispersion estimates by setting. For all panels, the colours of the estimate correspond to the model displayed.

The distribution of household sizes used in the model is displayed in Figure 17.

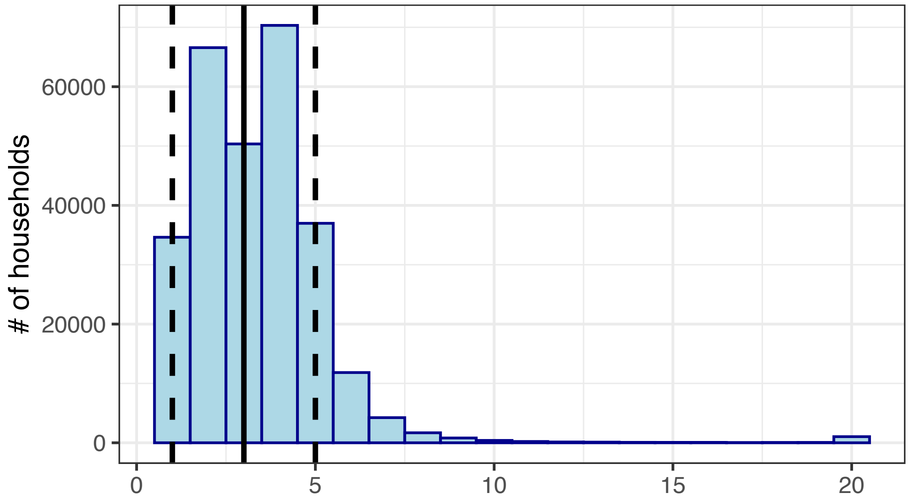


**Supplementary Figure 17.** Household size

##### *5.2.1. Coefficients of main model.*

**Supplementary Table 5.** Estimated coefficients, including 95% HDI, Markov Chain Standard Errors (MCSE), Effective Sample Size (ESS), R hat ($\hat{R}$) statistics. Please note that for risk factors, the raw coefficients are displayed and not IRRs.

| NPI | Risk factor | Setting | Group | Mean | HDI 2.5% | HDI 97.5% | MCSE mean | MCSE s.d. | ESS bulk | ESS tail | $\hat{R}$ |
|---|---|---|---|---|---|---|---|---|---|---|---|
| - | Overdispersion | - | Household | 38.435 | 31.285 | 47.326 | 0.132 | 0.104 | 989 | 870 | 1 |
| - | Overdispersion | - | Family (n/h) | 0.175 | 0.042 | 0.371 | 0.003 | 0.005 | 951 | 904 | 1 |
| - | Overdispersion | - | Community | 0.322 | 0.318 | 0.326 | 0 | 0 | 1016 | 872 | 1.01 |
| - | Overdispersion | - | School | 0.047 | 0.042 | 0.055 | 0 | 0 | 1013 | 960 | 1 |
| - | Overdispersion | - | Workplace | 0.094 | 0.086 | 0.106 | 0 | 0 | 977 | 997 | 1 |
| School restrictions | - | Household | - | 0.414 | 0.309 | 0.519 | 0.002 | 0.001 | 1093 | 997 | 1 |
| Workplace restrictions | - | Household | - | 0.354 | 0.268 | 0.447 | 0.001 | 0.001 | 1019 | 928 | 1 |
| Restriction on gathering size | - | Household | - | -0.964 | -1.042 | -0.884 | 0.001 | 0.001 | 1051 | 901 | 1 |
| Stay at home order | - | Household | - | 0.899 | 0.756 | 1.045 | 0.002 | 0.002 | 1041 | 955 | 1 |
| Restrictions on international travel | - | Household | - | 1.269 | 1.128 | 1.401 | 0.002 | 0.002 | 1082 | 813 | 1.01 |
| Facial coverings | - | Household | - | -0.036 | -0.21 | 0.149 | 0.003 | 0.002 | 1020 | 992 | 1 |
| - | Age | - | >60yrs | 0 | 0 | 0 | 0 | | 1000 | 1000 | |
| - | Age | - | 40-59yrs | 0.007 | -0.016 | 0.028 | 0 | 0 | 1032 | 1065 | 1 |
| - | Age | - | 19-39yrs | -0.014 | -0.037 | 0.008 | 0 | 0 | 1049 | 997 | 1 |
| - | Age | - | 11-18yrs | -0.166 | -0.192 | -0.141 | 0 | 0 | 916 | 919 | 1.01 |
| - | Age | - | 0-10yrs | -0.262 | -0.29 | -0.234 | 0 | 0 | 994 | 887 | 1 |
| School restrictions | - | Family (n/h) | - | -0.032 | -0.365 | 0.393 | 0.006 | 0.004 | 898 | 984 | 1.01 |
| Workplace restrictions | - | Family (n/h) | - | -0.062 | -0.347 | 0.238 | 0.005 | 0.004 | 983 | 999 | 1 |
| Restriction on gathering size | - | Family (n/h) | - | -0.333 | -0.634 | -0.042 | 0.005 | 0.004 | 991 | 988 | 1 |
| Stay at home order | - | Family (n/h) | - | 1.941 | 1.327 | 2.545 | 0.01 | 0.007 | 928 | 955 | 1 |
| Restrictions on international travel | - | Family (n/h) | - | 0.767 | 0.301 | 1.268 | 0.008 | 0.005 | 942 | 899 | 1 |
| Facial coverings | - | Family (n/h) | - | 3.065 | 2.489 | 3.563 | 0.009 | 0.006 | 856 | 996 | 1.01 |
| School restrictions | - | Community | - | -1.115 | -1.198 | -1.038 | 0.001 | 0.001 | 1035 | 855 | 1 |
| Workplace restrictions | - | Community | - | 0.229 | 0.161 | 0.287 | 0.001 | 0.001 | 1103 | 997 | 1 |
| Restriction on gathering size | - | Community | - | -0.461 | -0.518 | -0.4 | 0.001 | 0.001 | 1038 | 1042 | 1 |
| Stay at home order | - | Community | - | 0.642 | 0.523 | 0.758 | 0.002 | 0.001 | 1023 | 917 | 1 |
| Restrictions on international travel | - | Community | - | 0.213 | 0.11 | 0.318 | 0.002 | 0.001 | 819 | 782 | 1.01 |
| Facial coverings | - | Community | - | -1.199 | -1.312 | -1.082 | 0.002 | 0.001 | 987 | 946 | 1 |
| School restrictions | - | School | - | -0.099 | -0.315 | 0.113 | 0.003 | 0.003 | 1006 | 953 | 1 |
| Workplace restrictions | - | School | - | 0.463 | 0.276 | 0.643 | 0.003 | 0.002 | 1048 | 955 | 1 |
| Restriction on gathering size | - | School | - | -0.946 | -1.101 | -0.758 | 0.003 | 0.002 | 1092 | 952 | 1 |
| Stay at home order | - | School | - | -0.07 | -0.383 | 0.239 | 0.005 | 0.004 | 1014 | 885 | 1.01 |
| Restrictions on international travel | - | School | - | -0.245 | -0.542 | 0.072 | 0.005 | 0.003 | 1011 | 989 | 1 |
| Facial coverings | - | School | - | 0.541 | 0.219 | 0.888 | 0.005 | 0.004 | 1035 | 1041 | 1 |

| | | | | | | | | | | | |
|---|---|---|---|---|---|---|---|---|---|---|---|
| - | Vaccination | - | Unvaccinated | 0 | 0 | 0 | 0 | | 1000 | 1000 | |
| - | Vaccination | - | One Dose | -0.155 | -0.182 | -0.131 | 0 | 0 | 874 | 997 | 1.01 |
| - | Vaccination | - | Two Dose | -0.235 | -0.252 | -0.215 | 0 | 0 | 883 | 937 | 1 |
| - | Variant | - | B.1.177 | 0 | 0 | 0 | 0 | | 1000 | 1000 | |
| - | Variant | - | Alpha | 0.123 | 0.1 | 0.149 | 0 | 0 | 965 | 939 | 1 |
| - | Variant | - | Delta | 0.024 | -0.014 | 0.066 | 0.001 | 0 | 1103 | 1090 | 1 |
| - | Variant | - | Eta | 0.028 | -0.094 | 0.148 | 0.002 | 0.001 | 1064 | 1041 | 1 |
| - | Variant | - | Omicron | 0.257 | 0.205 | 0.302 | 0.001 | 0.001 | 1091 | 955 | 1 |
| School restrictions | - | Workplace | - | 0.481 | 0.308 | 0.65 | 0.003 | 0.002 | 924 | 1024 | 1 |
| Workplace restrictions | - | Workplace | - | 0.06 | -0.088 | 0.189 | 0.002 | 0.002 | 1109 | 957 | 1 |
| Restriction on gathering size | - | Workplace | - | 0.248 | 0.115 | 0.386 | 0.002 | 0.002 | 1217 | 952 | 1 |
| Stay at home order | - | Workplace | - | 0.505 | 0.28 | 0.777 | 0.004 | 0.003 | 1013 | 996 | 1 |
| Restrictions on international travel | - | Workplace | - | 0.593 | 0.337 | 0.831 | 0.004 | 0.003 | 1100 | 1026 | 1 |
| Facial coverings | - | Workplace | - | -2.355 | -2.748 | -2 | 0.006 | 0.005 | 904 | 949 | 1 |
| - | - | Household | Nordjydanmark | 0.048 | -0.057 | 0.166 | 0.002 | 0.003 | 1061 | 997 | 1 |
| - | - | Household | Midjydanmark | -0.012 | -0.144 | 0.08 | 0.002 | 0.004 | 1083 | 988 | 1 |
| - | - | Household | Syddanmark | 0.066 | -0.051 | 0.166 | 0.002 | 0.004 | 1020 | 997 | 1 |
| - | - | Household | Hovedstaden | -0.056 | -0.154 | 0.067 | 0.002 | 0.004 | 1055 | 1040 | 1 |
| - | - | Household | Sjaelland | -0.062 | -0.177 | 0.042 | 0.002 | 0.004 | 1011 | 1088 | 1 |
| - | - | Family (n/h) | Nordjydanmark | -0.066 | -0.26 | 0.09 | 0.004 | 0.007 | 923 | 913 | 1 |
| - | - | Family (n/h) | Midjydanmark | -0.057 | -0.228 | 0.088 | 0.003 | 0.006 | 957 | 915 | 1 |
| - | - | Family (n/h) | Syddanmark | 0.055 | -0.073 | 0.223 | 0.003 | 0.005 | 750 | 919 | 1 |
| - | - | Family (n/h) | Hovedstaden | -0.018 | -0.154 | 0.116 | 0.003 | 0.007 | 942 | 918 | 1 |
| - | - | Family (n/h) | Sjaelland | 0.049 | -0.087 | 0.239 | 0.003 | 0.005 | 954 | 954 | 1.01 |
| - | - | Community | Nordjydanmark | -0.036 | -0.107 | 0.054 | 0.001 | 0.001 | 960 | 863 | 1 |
| - | - | Community | Midjydanmark | 0.003 | -0.077 | 0.093 | 0.001 | 0.001 | 944 | 886 | 1 |
| - | - | Community | Syddanmark | -0.028 | -0.119 | 0.043 | 0.001 | 0.001 | 932 | 852 | 1 |
| - | - | Community | Hovedstaden | 0.087 | 0.005 | 0.167 | 0.001 | 0.001 | 904 | 785 | 1 |
| - | - | Community | Sjaelland | -0.027 | -0.113 | 0.054 | 0.001 | 0.001 | 897 | 884 | 1.01 |
| - | - | School | Nordjylland | 0.136 | -0.108 | 0.367 | 0.004 | 0.005 | 1012 | 877 | 1 |
| - | - | School | Midjylland | 0.138 | -0.076 | 0.356 | 0.004 | 0.005 | 930 | 775 | 1 |
| - | - | School | Syddanmark | -0.032 | -0.263 | 0.192 | 0.004 | 0.005 | 964 | 943 | 1 |
| - | - | School | Hovedstaden | -0.125 | -0.349 | 0.084 | 0.004 | 0.005 | 977 | 969 | 1 |
| - | - | School | Sjaelland | -0.155 | -0.385 | 0.064 | 0.004 | 0.005 | 990 | 834 | 1 |
| - | - | Workplace | Nordjylland | 0.102 | -0.005 | 0.242 | 0.002 | 0.002 | 976 | 958 | 1 |
| - | - | Workplace | Midjylland | -0.046 | -0.164 | 0.076 | 0.002 | 0.003 | 1127 | 1018 | 1 |
| - | - | Workplace | Syddanmark | -0.076 | -0.208 | 0.03 | 0.002 | 0.003 | 1026 | 1016 | 1 |
| - | - | Workplace | Hovedstaden | 0.026 | -0.096 | 0.135 | 0.002 | 0.003 | 1004 | 958 | 1 |
| - | - | Workplace | Sjaelland | -0.02 | -0.131 | 0.105 | 0.002 | 0.003 | 1025 | 1089 | 1 |
| - | Holiday | - | Household | -0.225 | -0.259 | -0.196 | 0 | 0 | 1112 | 1039 | 1 |
| - | Holiday | - | Family (n/h) | 0.095 | -0.018 | 0.204 | 0.002 | 0.001 | 1011 | 933 | 1 |
| - | Holiday | - | Community | -0.006 | -0.028 | 0.015 | 0 | 0 | 938 | 922 | 1 |
| - | Holiday | - | School | -0.769 | -0.846 | -0.697 | 0.001 | 0.001 | 940 | 997 | 1 |
| - | Holiday | - | Workplace | -0.2 | -0.257 | -0.142 | 0.001 | 0.001 | 1120 | 880 | 1 |
| - | Hurdle rate | - | Household | 0.538 | 0.521 | 0.557 | 0 | 0 | 946 | 981 | 1 |
| - | Hurdle rate | - | Family (n/h) | 0.635 | 0.255 | 0.89 | 0.006 | 0.005 | 950 | 876 | 1 |
| - | Hurdle rate | - | Community | 0 | 0 | 0.001 | 0 | 0 | 1100 | 1034 | 1 |
| - | Hurdle rate | - | School | 0.078 | 0 | 0.211 | 0.002 | 0.002 | 995 | 993 | 1 |
| - | Hurdle rate | - | Workplace | 0.046 | 0 | 0.134 | 0.001 | 0.001 | 806 | 956 | 1 |

#### 5.3. Sensitivity Analysis

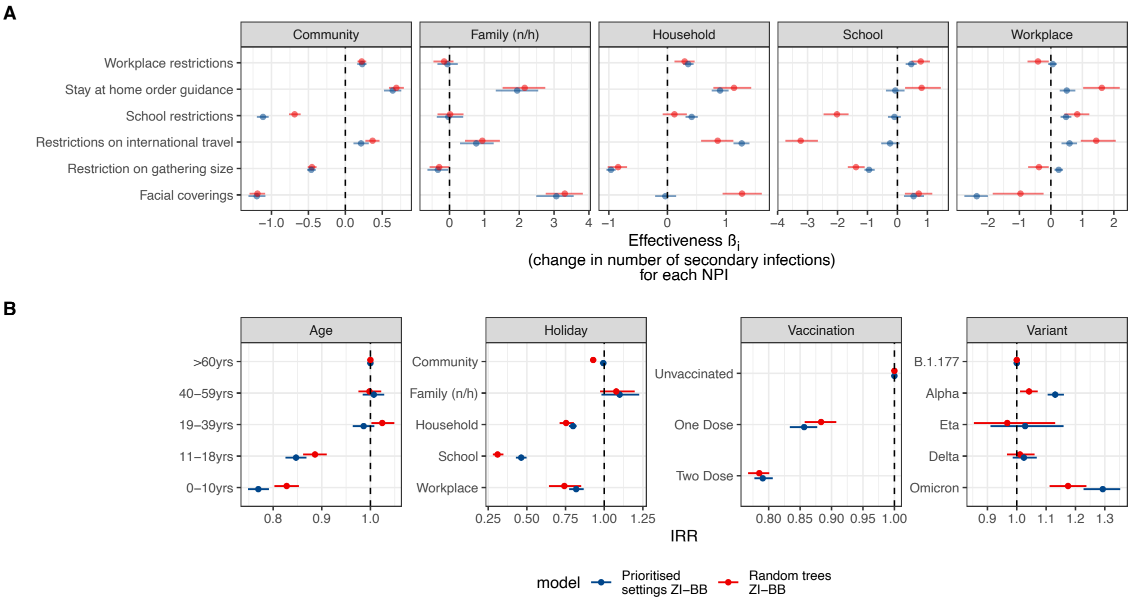


**Supplementary Figure 18.** Effectiveness of NPIs prioritised settings vs random trees sensitivity analysis: (A) Effect sizes of association between number of infections caused by an infector and level of each NPI by setting. (B) Risk heterogeneities of the number of secondary infections by Age, School holidays, Vaccination status and SARS-CoV-2 variant. For both panels, the colours of the estimate correspond to the model displayed. Family (n/h) = Family not living in the same household as the infector.

In this section, we conduct several sensitivity analyses to examine the robustness of our estimates in response to changes in the underlying transmission chains.

***5.3.1. Random vs Prioritised Networks.*** We describe the construction of the transmission chains, both based on prioritising the settings information of an infector/infectee pair and randomly choosing amongst all potential infectors. In the former case, we sample for each individual an infector from only those plausible infectors who share a setting with the individual. Under this sampling method, we prioritise selecting an infector from settings in the following order: households, then schools, then workplaces, then family members. If an individual has multiple plausible infectors from the same setting, then we choose randomly from these infectors. If an individual has no plausible infectors in any of these settings, then we sample an infector randomly from the set of all of their plausible infectors, irrespective of setting. We refer to plausible transmission trees derived in this way as "prioritised settings" trees.

We refer to plausible transmission trees derived by randomly choosing an infector for each individual from their total set of plausible infectors as "random trees", and these trees have an underlying assumption of homogeneous mixing, such that only the edge weights in $G$ are taken into consideration in the sampling of transmission pairs. In contrast, the prioritised setting trees assume heterogeneity in mixing based on individuals' social network connections, with transmission pairs belonging to the same setting selected preferentially.

In Figure 18, we show the results of the association of NPIs and transmission by setting in panel (A) and risk heterogeneities in panel (B). It is important to recall that in transmission chains based on random selection of infectors, there are significantly fewer secondary infections in non-community settings.

The results for the community setting are broadly similar, although the association between school restrictions and transmission is weaker in magnitude. Regarding restrictions on international travel, we observed a positive association with transmission. The household setting also displayed consistent results, except that face mask restrictions were more positively associated with transmission. The other three settings each had only a small number of secondary infections (see Figure 7).

The results on risk heterogeneities were broadly consistent. For age, the results based on random trees showed less difference in risk for the 19-39 year old and 40-59 year old age groups; however, this is inconsistent with cohort studies such as Goldstein et al. [30]. The results for vaccine effectiveness against transmission showed lower VE for both one and two doses for the model using random trees relative to the prioritised settings. For one dose, the VE estimate for the random tree-based model was 12.5% (94% HDI: 10.3% to 14.8%) versus 15.5% (94% HDI: 13.1% to 18.2%) for the prioritised settings. For two doses, the VE estimate for the random tree-based model was 22.2% (94% HDI: 20.6% to 23.9%) versus 25.1% (94% HDI: 21.5% to 25.2%) for the prioritised settings.

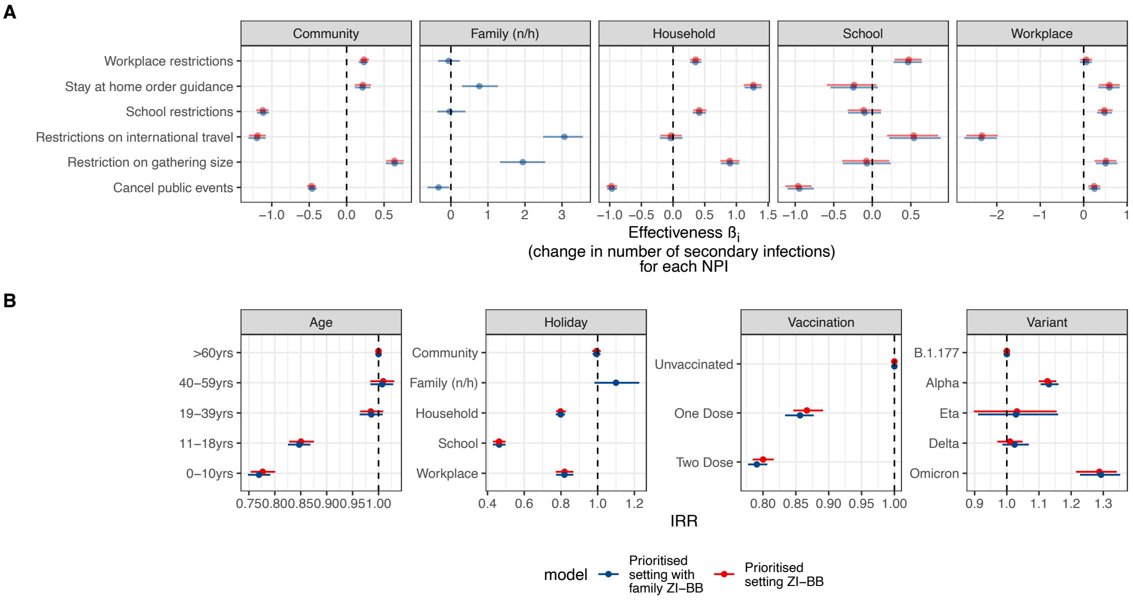


**Supplementary Figure 19.** Effectiveness of NPIs inclusion of non-household family sensitivity analysis: (A) Effect sizes of association between number of infections caused by an infector and level of each NPI by setting. (B) Risk heterogeneities of the number of secondary infections by Age, School holidays, Vaccination status and SARS-CoV-2 variant. For both panels, the colours of the estimate correspond to the model displayed. Family (n/h) = Family not living in the same household as the infector.

***5.3.2. Prioritised settings with and without 'Family (outside household)'.*** The number of infector/infectee pairs in the family outside the household setting is small relative to all other settings (Figure 19). We included the setting in the model because we were interested in the associations between restrictions and transmissions in that setting, especially in comparison to the household setting. The comparison of the results showed that the inclusion of the family outside the household setting had no impact on the other estimates.

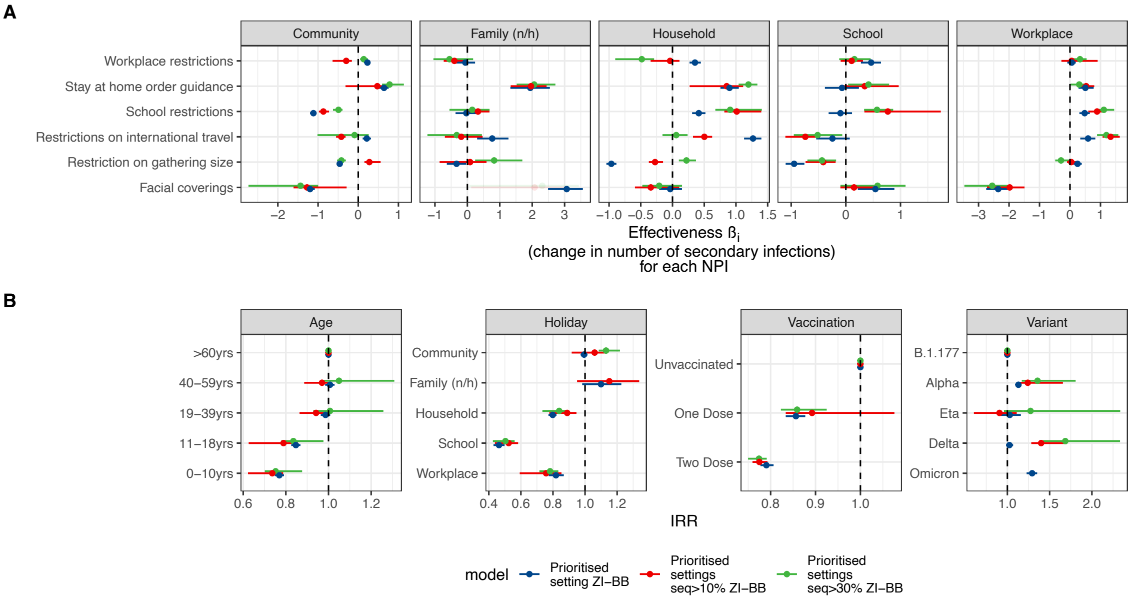


**Supplementary Figure 20.** Effectiveness of NPIs level of sequencing sensitivity analysis: (A) Effect sizes of association between number of infections caused by an infector and level of each NPI by setting. (B) Risk heterogeneities of the number of secondary infections by Age, School holidays, Vaccination status and SARS-CoV-2 variant. For both panels, the colours of the estimate correspond to the model displayed. Family (n/h) = Family not living in the same household as the infector.

***5.3.3. Prioritised settings with different levels of sequencing required.*** The proportion of infections sequenced varied over the pandemic (Figure 1). In Figure 20, we show the different estimates depending on the level of sequencing. The blue estimates correspond to the whole study period, the red estimates correspond to the period with at least 10% of infections sequenced, and the green estimates correspond to the period with at least 30% of infections sequenced (and hence is the shortest of the three). It is worth noting that the estimates overall are consistent, varying only in their magnitude. No estimates for the Omicron variant are shown for the 10% and 30% minimum sequencing models, as neither included data from the Omicron period.

***5.3.4. Prioritised settings comparison of Maximum Likelihood trees vs random trees.*** One further comparison is between transmission trees, which are chosen at random, versus the maximum likelihood tree for that specific setup. In Figure 21, we show the results for three different prioritised setting trees (all three chosen at random) and the maximum likelihood tree. The association between the level of NPIs and transmission is broadly consistent across community, family outside the household, household, and workplace settings. For schools, we observe more differences between the estimates, with the maximum likelihood results indicating significant effects for face coverings, protection of the elderly, school restrictions, and stay-at-home orders. In contrast, the random selection-based results indicate results close to zero.

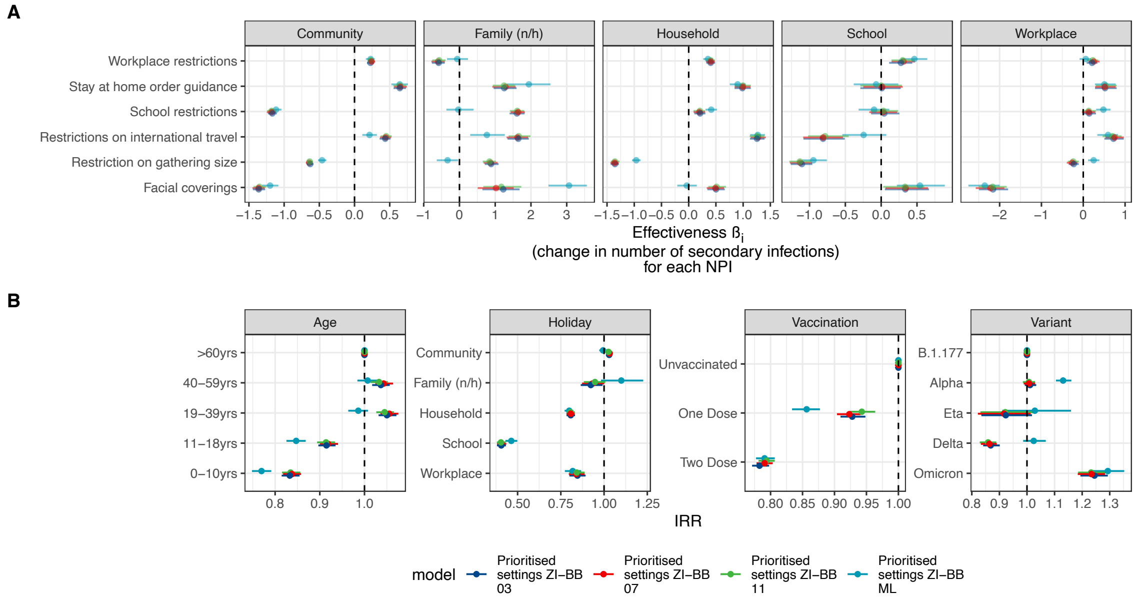


**Supplementary Figure 21.** Effectiveness of NPIs ML vs individual trees sensitivity analysis: (A) Effect sizes of association between number of infections caused by an infector and level of each NPI by setting. (B) Risk heterogeneities of the number of secondary infections by Age, School holidays, Vaccination status and SARS-CoV-2 variant. For both panels, the colours of the estimate correspond to the model displayed. Family (n/h) = Family not living in the same household as the infector.

Considering risk heterogeneities, we observed a smaller magnitude of effect size for the 40-59 year old group, with all other age groups showing very similar magnitudes of effect size. The results for the vaccination effectiveness with regard to transmission are lower for the random selection of trees relative to the maximum likelihood tree.

### Appendix 6: List of members of the Danish COVID-19 Genome Consortium (DCGC)

Below is a list of members of The Danish COVID-19 Genome Consortium. Individuals may appear both as authors and as acknowledged DCGC members.

**Anders Fomsgaard[1], Morten Rasmussen[1], Søren M. Karst[1], Jannik Fonager[1], Vithiagaran Gunalan[1], Marc Bennedbæk[1], Aleksander Ring[1], Marc Stegger[1], Raphael Sieber[1], Kirsten Ellegaard[1], Anna C. Ingham[1], Thor B. Johannesen[1], Gitte Nygaard Aasbjerg[1], Berit Lilje[1], Kim L. Ng[1], Sofie M. Edslev[1], Sharmin Baig[1], Povilas Matusevicius[1], Lars Bustamante Christoffersen[1], Man-Hung Eric Tang[1], Leandro Andrés Escobar-Herrera[1], Casper Westergaard[1], Christina Wiid Svarrer[1], Nour Saad Al-Tamimi[1], Marie Bækvad-Hansen[1], Jonas Byberg-Grauholm[1], Mette Theilgaard Christiansen[1], Karen Marie Jørgensen[2], Tobias Nikolaj Gress Hansen[2], Esben Mørk Hartmann[2], Nicolai Balle Larsen[2], Arieh Cohen[2], Kasper Holtz Uhre[3], Christian Nielsen[3], Anders Jensen[3], Jakob Matias Melbye[3], Bartlomiej Wilkowski[3], Karina Meden Sørensen[3], Mads Albertsen[4], Thomas Y. Michaelsen[4], Vang Le-Quy[4], Mantas Sereika[4], Rasmus H. Kirkegaard[4], Kasper S. Andersen[4], Martin H. Andersen[4], Karsten K. Hansen[4], Mads Boye[4], Mads P. Bach[4], Peter Dissing[4], Anton Drastrup-Fjordbak[4], Michael Collin[4], Finn Büttner[4], Jakob Brandt[4], Simon Knuttson[4], Emil A. Sørensen[4], Thomas B. N. Jensen[4], Trine Sørensen[4], Celine Petersen[4], Clarisse Chiche-Lapierre[4], Frederik T. Hansen[4], Emilio F. Collados[4], Amalie Berg[4], Susanne R. Bielidt[4], Sebastian M. Dall[4], Erika Dvarionaite[4], Susan H. Hansen[4], Vibeke R. Jørgensen[4], Trine B. Nicolajsen[4], Wagma Saei[4], Stine K. Østergaard[4], Martin Bøgsted[4], Rasmus Brøndu[4], Katja Hose[4], Tomer Sagi[4], Miroslav Pakanec[4], Henrik Krarup[5], David Fuglsang-Damgaard[5], Mette Mølvadgaard[5], Marc T. K. Nielsen[5], Kristian Schønning[6], Martin S. Pedersen[6], Rasmus L. Marvig[6], Nikolai Kirkby[6], Uffe V. Schneider[7], Jose A. S. Castruita[7], Nana G. Jacobsen[7], Christian Ø. Andersen[7], Mette Christiansen[8], Ole H. Larsen[8], Kristian A. Skipper[8], Søren Vang[8], Kurt J. Handberg[8], Carl M. Kobel[8], Camilla Andersen[8], Irene H. Tarpgaard[8], Svend Ellermann-Eriksen[8], Marianne Skov[9], Thomas V. Sydenham[9], Lene Nielsen[10], Line L. Nilsson[10], Martin B. Friis[10],Thomas Sundelin[10], Thomas A. Hansen[10], Dorte T. Andersen[11], John E. Coia[11], Anders Jensen[12], Ea S. Marmolin[12], Xiaohui C. Nielsen[13], Christian H. Schouw[13], Simon Knutsson[4], Marianne Kragh Thomsen[8], Tine Sneibjerg Ebsen[8], Morten Hoppe[10], Asta Laugesen[13], Laus Vejrum[13], Rikke Johansen[13], Josefine Møller[13], Tina Madsen[13]**

[1]Statens Serum Institut, Copenhagen, Denmark
[2]TestCenter Denmark, Copenhagen, Denmark
[3]Danish National Biobank, Copenhagen, Denmark
[4]Aalborg University, Aalborg, Denmark
[5]Aalborg University Hospital, Aalborg, Denmark
[6]Rigshospitalet, Copenhagen University Hospital, Copenhagen, Denmark
[7]Hvidovre Hospital, Hvidovre, Denmark
[8]Aarhus University Hospital, Aarhus, Denmark
[9]Odense University Hospital, Odense, Denmark
[10]Herlev Hospital, Herlev, Denmark
[11]Sydvestjysk Sygehus, Denmark
[12]Sygehus Lillebaelt, Denmark
[13]Zealand University Hospital, Denmark
